# Quantifying Large Language Model Influence in Brain Computer Interface Communication for Amyotrophic Lateral Sclerosis

**DOI:** 10.64898/2026.08.20.26360941

**Authors:** Alon Gorenshtein, Mahmud Omar, Eric Jia, Yosef Adiniaev, Oved Daniel, Jonathan Kruskal, Muneeb Ahmed, Olga Brook, Yiftach Barash, Eyal Klang

## Abstract

Large language models are integrated into brain-computer interfaces for communication, but accuracy does not show whether an emitted character depended on neural evidence or on the language-model prior. We retrospectively re-decoded 3,373 P300-speller selections from 47 people with amyotrophic lateral sclerosis, reconstructing a neural posterior and combining it with 25 language priors ranging from 5-grams to 46.7-billion-parameter models. Under held-out, per-source calibration and equal fusion weighting, the prior accounted for a participant-weighted mean 8.6% of posterior displacement (median across selections, 2.9%); the corresponding neural contribution fraction was 0.914 (95% CI, 0.896-0.934). In 4.4% of selections (95% CI, 3.5-5.3), the fused system emitted the intended character although neural evidence alone would not have selected it. Results were similar across 21 neural language models. Accuracy alone does not reveal how strongly a fused brain-computer interface decision depends on its language prior.

## Introduction

Large language models are increasingly being integrated into brain-computer interfaces that restore communication for people who have lost speech and movement to amyotrophic lateral sclerosis. The P300 speller is one of the most extensively studied non-invasive systems,^1^ alongside approaches extending to implanted electrocorticography.^2^ Because each selection carries only a few bits of neural information, spellers have long combined neural evidence with a language prior that narrows the candidate set.^3,4^ Current language models provide richer, context-sensitive priors and have been evaluated in simulated and online spellers.^5–7^ Recent work has reported near-ceiling fused performance and argued that neural decoding, rather than language modelling, is now the main bottleneck.^8^

That conclusion raises a question accuracy and information-transfer rate cannot answer: how much did each emitted character depend on neural evidence, and how much on the language prior? Speier and colleagues showed that prior information can inflate reported communication rates and proposed a mutual-information correction.^9^ Their measure operates on the completed output string, not on individual selections, because the necessary per-selection data were then unavailable. Raw legacy recordings, per-symbol stimulus channels, and online feedback streams are now public,^10^ while neural language models increasingly supply the prior. Users of augmentative and alternative communication and their clinicians have also raised questions of authorship and agency in model-assisted output.^11,12^ Information-theoretic measures of human contribution to AI-assisted text exist,^13^ but have not been applied at the character level in a brain-computer interface.

We therefore asked two related questions: how strongly does each emitted character depend on neural evidence rather than the language-model prior, and how often does the fused system emit the intended character when neural evidence alone would have selected another? We estimated both in a retrospective re-decoding study of archived online P300-speller sessions from people with ALS.

## Results

### Cohort

The analysis included 3,373 online selections from 47 participants with ALS across 115 sessions and four source studies (Table 1). Participants contributed a median of 84 selections (range 4-154), spelling 289 distinct words, including 28 digit strings. Median calibration-decoder AUC was 0.795 (IQR 0.686-0.861). The offline neural posterior matched the archive’s online selection in 69.6% of trials (chance 2.8%) and was correct against the intended character in 71.8%, compared with 81.9% online accuracy.

**Table 1.**
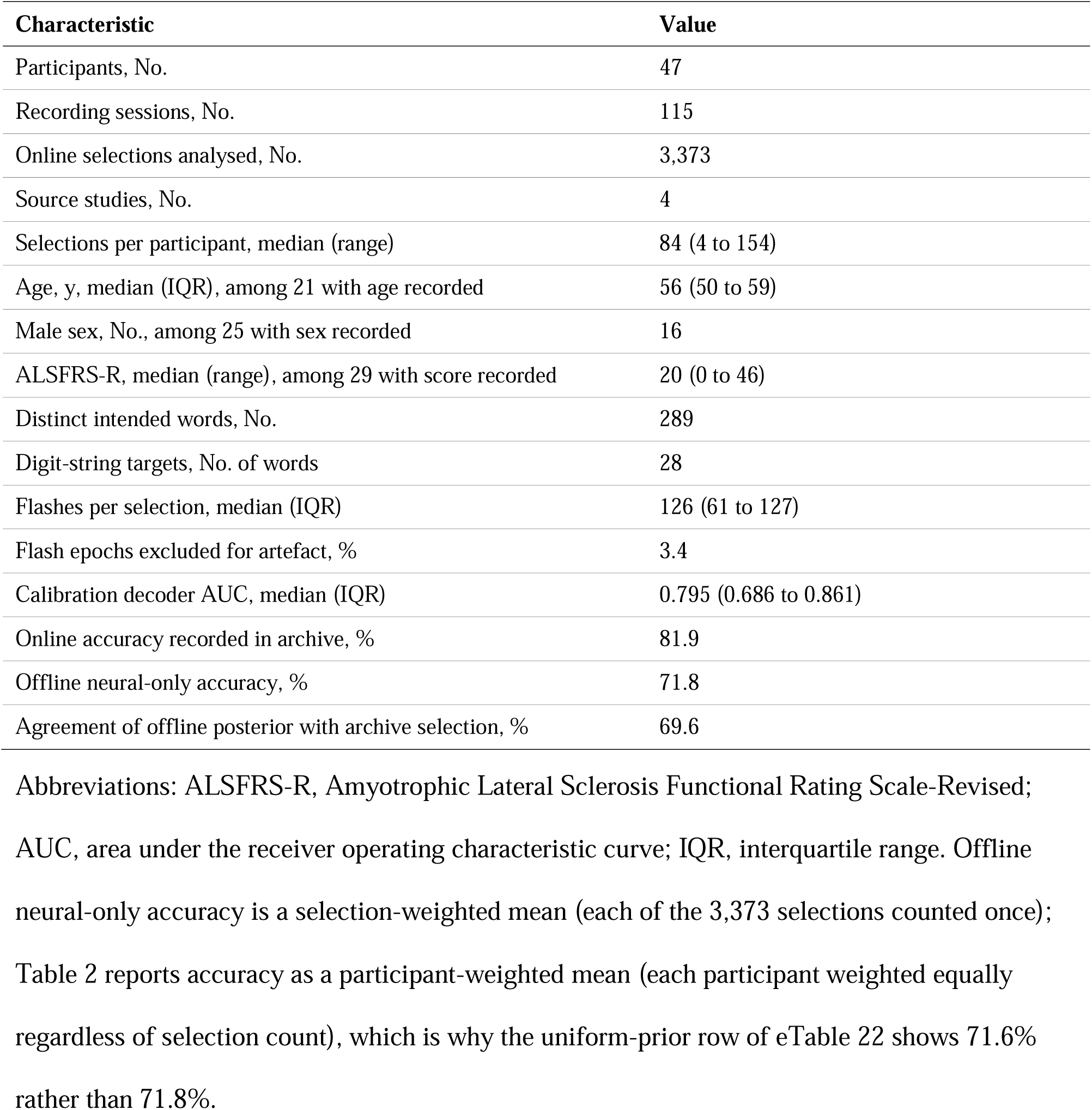
Cohort characteristics.

| Characteristic | Value |
| --- | --- |
| Participants, No. | 47 |
| Recording sessions, No. | 115 |
| Online selections analysed, No. | 3,373 |
| Source studies, No. | 4 |
| Selections per participant, median (range) | 84 (4 to 154) |
| Age, y, median (IQR), among 21 with age recorded | 56 (50 to 59) |
| Male sex, No., among 25 with sex recorded | 16 |
| ALSFRS-R, median (range), among 29 with score recorded | 20 (0 to 46) |
| Distinct intended words, No. | 289 |
| Digit-string targets, No. of words | 28 |
| Flashes per selection, median (IQR) | 126 (61 to 127) |
| Flash epochs excluded for artefact, % | 3.4 |
| Calibration decoder AUC, median (IQR) | 0.795 (0.686 to 0.861) |
| Online accuracy recorded in archive, % | 81.9 |
| Offline neural-only accuracy, % | 71.8 |
| Agreement of offline posterior with archive selection, % | 69.6 |

Abbreviations: ALSFRS-R, Amyotrophic Lateral Sclerosis Functional Rating Scale-Revised; AUC, area under the receiver operating characteristic curve; IQR, interquartile range. Offline neural-only accuracy is a selection-weighted mean (each of the 3,373 selections counted once); Table 2 reports accuracy as a participant-weighted mean (each participant weighted equally regardless of selection count), which is why the uniform-prior row of eTable 22 shows 71.6% rather than 71.8%.

### Co-primary outcomes

#### Under the primary held-out calibrated fusion, the prior accounted for a participant-weighted mean 8.6% of posterior displacement

The distribution was strongly right-skewed (unweighted median 2.9%): most selections were almost entirely neural, while a minority depended substantially on the prior. Equivalently, the neural contribution fraction was 0.914 (95% CI 0.896-0.934). The raw, uncalibrated distribution was even more concentrated near zero (median prior share 0.1%; eFigure 3, eTable 14). Because any non-uniform prior mechanically moves the fused posterior away from the neural posterior, a context-permutation control tested whether that movement reflected selection-specific linguistic context rather than generic posterior sharpening (Sensitivity analyses).

**Table 2.**
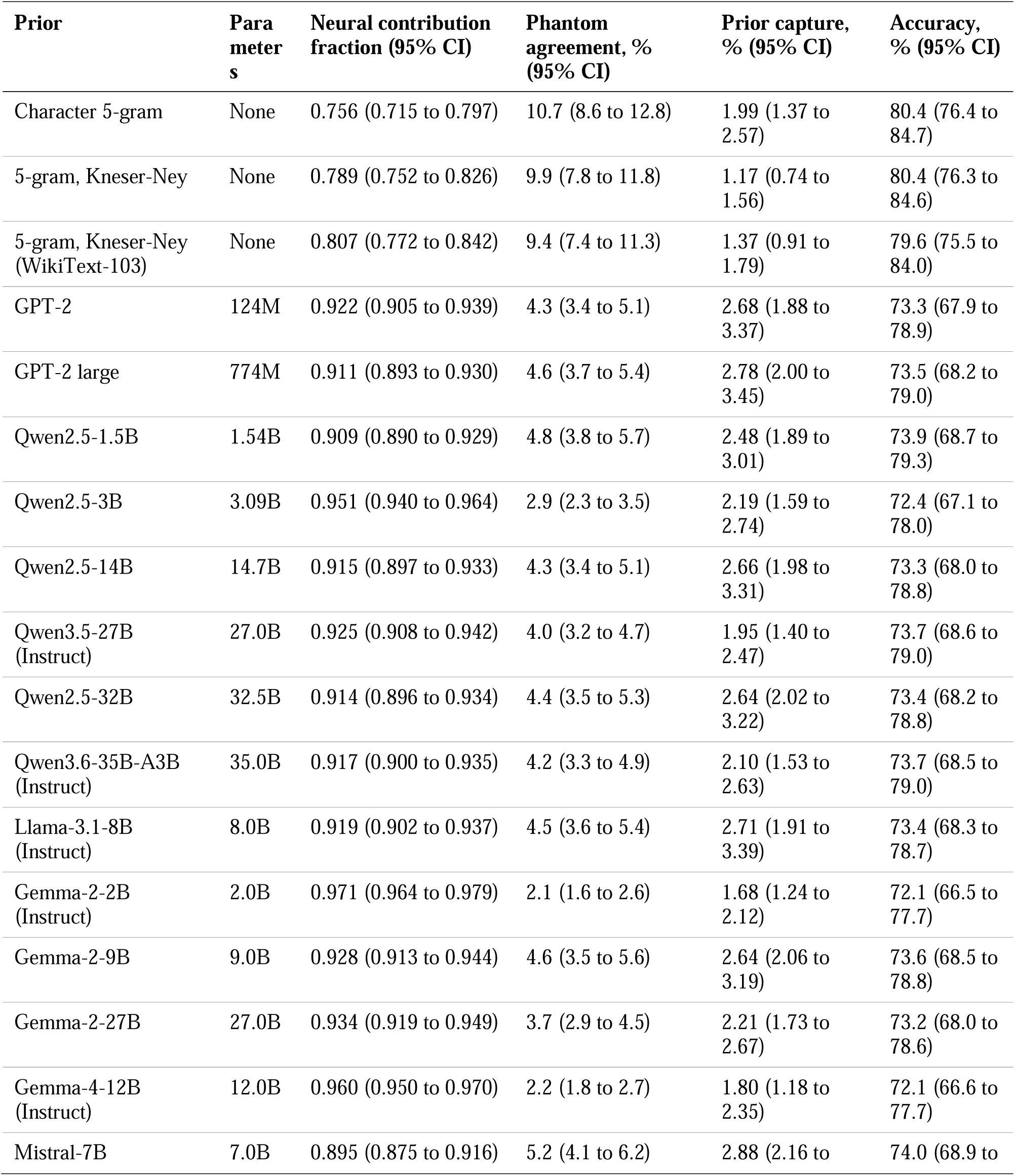
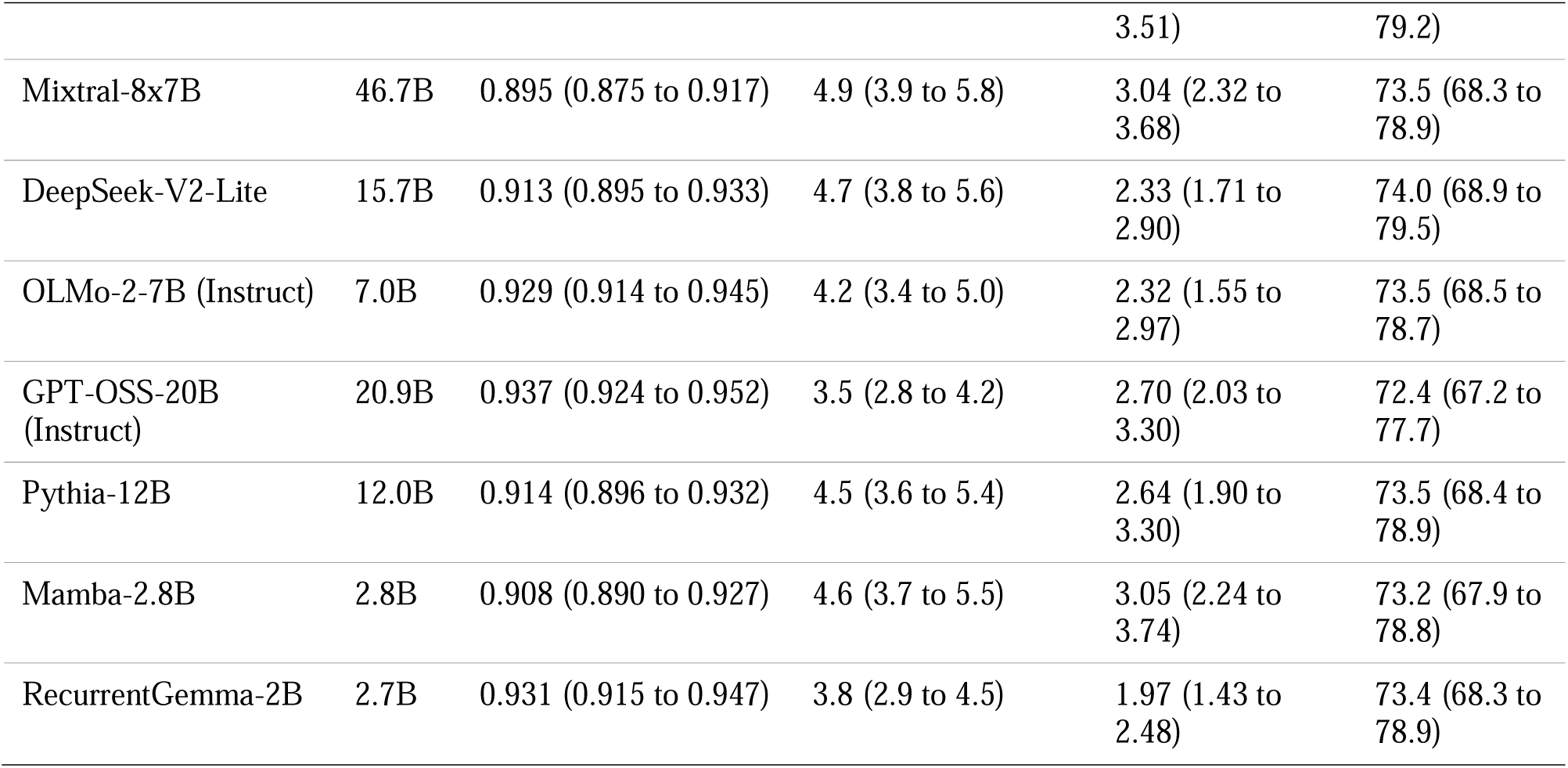
Attribution measures across the language-model prior ladder. Table 2 reports the full prior ladder after held-out, per-source temperature calibration; eTable 22 reports raw, uncalibrated fusion. The uniform prior is omitted because temperature scaling cannot change a uniform distribution and appears only in eTable 22. Prior capture was calculated separately for each prior. The Qwen2.5-32B row is the reference-prior estimate reported in the primary results.

#### Phantom agreement occurred in 4.4% of selections (95% CI 3.5-5.3), about 1 in 23

This outcome denotes a correct fused character that neural evidence alone would not have selected. The estimate met the prespecified co-primary threshold, so secondary outcomes were examined.

Across 21 neural language models, median estimates were similar (neural contribution fraction 0.919; phantom agreement 4.3%).

Among 166 phantom-agreement selections under calibrated fusion, the intended character was already the neural posterior’s runner-up in 98 (59.0%) and third-ranked in 35 (21.1%); it ranked fourth or lower in 29 (17.5%) and tied for first in 4 (2.4%) (Figure 4; eTable 26). Median neural probability on the intended character was 0.16 (IQR 0.10-0.24), and the median margin behind the neural top choice was 0.11 (0.04-0.21). Most were therefore close neural decisions tipped by the prior, although a small minority showed stronger conflict (Figure 4D; raw fusion in eTable 11).

#### An emitted-context sensitivity analysis produced similar estimates

When each new character was conditioned on earlier fused emissions rather than the intended prefix, phantom agreement decreased by 0.7 percentage points (95% CI 0.1-1.4) and the neural contribution fraction by 1.9 points (1.3-2.5; eTable 27). Emitted and intended context diverged in 35.7% of selections (1,203 of 3,373), so intended-context analysis was not uniformly an upper bound.

### Secondary outcomes

#### The prior overturned a correct neural choice in 2.6% of selections (95% CI 2.0-3.2), about 1 in 38

Neural override, a correct fused emission that the prior alone would not have selected, occurred in 62.6% (57.3-67.9%). Adding the prior increased accuracy by 1.8 percentage points (1.0-2.5). Raw-fusion estimates were 1.8%, 63.4%, and 2.0 points, respectively.

#### Model influence increased as linguistic context accumulated within a word

Phantom agreement rose from 2.5% at the first character to 7.7% at the sixth, while the neural contribution fraction fell from 0.934 to 0.879 (Figure 2A; eTable 28). Each additional character was associated with 1.19-fold higher odds of phantom agreement (95% CI 1.07-1.32; *P*=.001; adjusted *P*=.002). The bounded model predicted a 0.039 decrease in neural contribution across the six positions (0.027-0.050; *P*<.001; linear mixed-model slope -0.0081 per character, adjusted *P*<.001; eTable 30).

**Figure 1.**
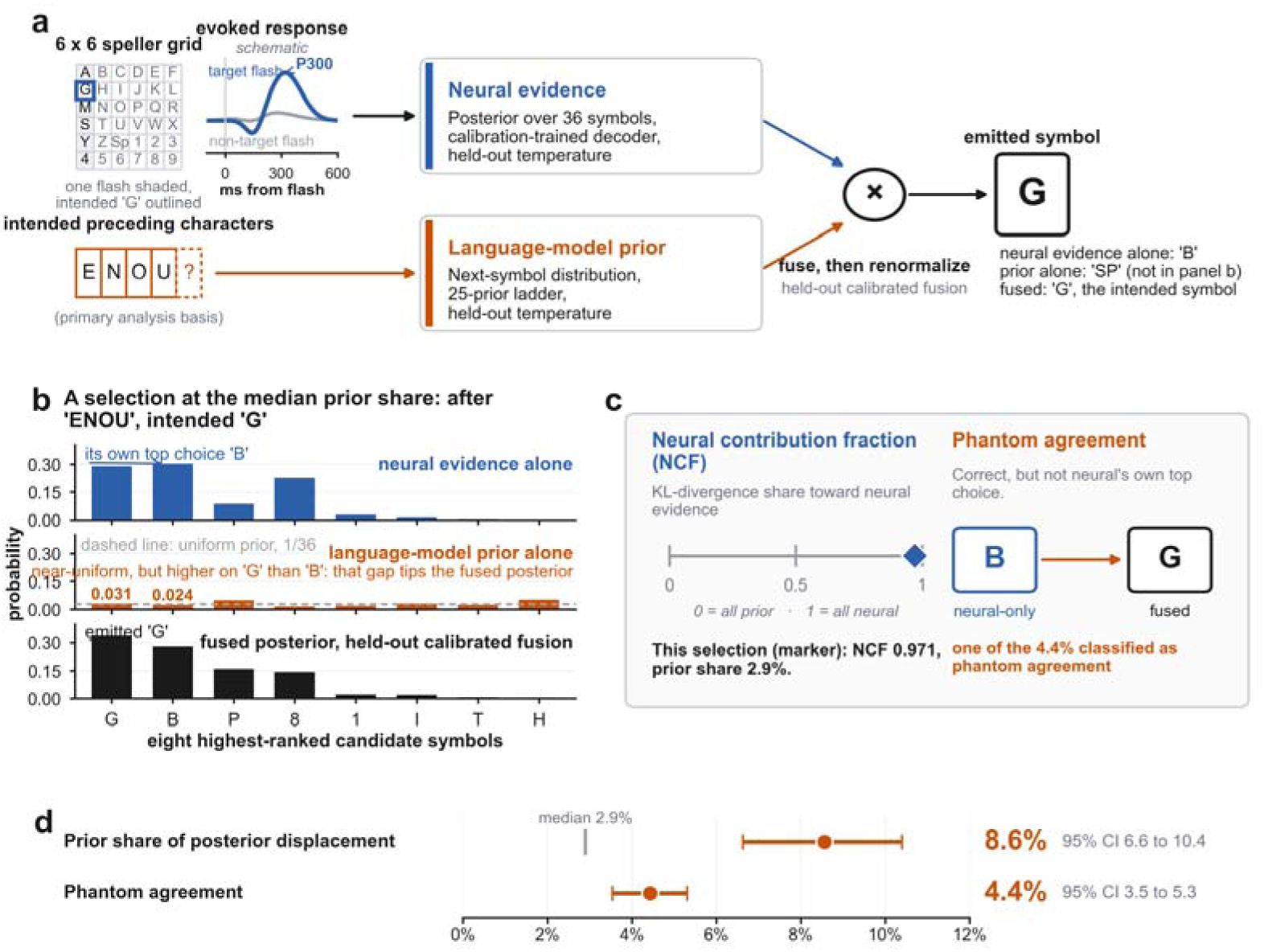
From flashed grid to emitted character: how each selection was attributed between the participant’s neural evidence and the language-model prior. (a) The reconstruction pipeline. Flashes of the 36-symbol grid evoke responses that a calibration-trained decoder accumulates into a posterior over the alphabet; the language model, conditioned on the intended preceding characters, supplies a prior over the next symbol; each source is then rescaled by a temperature fitted on held-out participant folds, and the two are multiplied and renormalized, the fused posterior’s highest-probability symbol being the emitted character. The grid is drawn with one flash shown as an illuminated column, although checkerboard conditions instead illuminate arbitrary symbol subsets, and the evoked-response trace is schematic rather than averaged data. (b) One selection, chosen by a rule fixed in advance as the selection whose prior share of posterior displacement lies closest to the cohort median among those with at least four preceding characters: the neural posterior, the prior, and the fused posterior across the eight highest-ranked candidate symbols, all under the same held-out calibrated fusion. The neural posterior’s own highest probability fell on B; the prior, close to uniform after calibration, nonetheless placed more probability on the intended G than on B, and that difference, annotated on the middle panel because it is too small to read off bars drawn on the shared scale, was enough to raise G above B in the fused posterior. (c) The two per-selection measures, with this selection’s values. (d) Headline co-primary estimates, each drawn as a point estimate with a bar spanning its 95% confidence interval on a shared percentage axis: the calibrated neural contribution fraction’s complement (the prior’s mean share of posterior displacement) and phantom agreement, both under held-out calibrated fusion with oracle context. The gray tick on the upper row marks the median prior share across selections. Intervals are from 2,000 participant-cluster bootstrap replicates.

**Figure 2.**
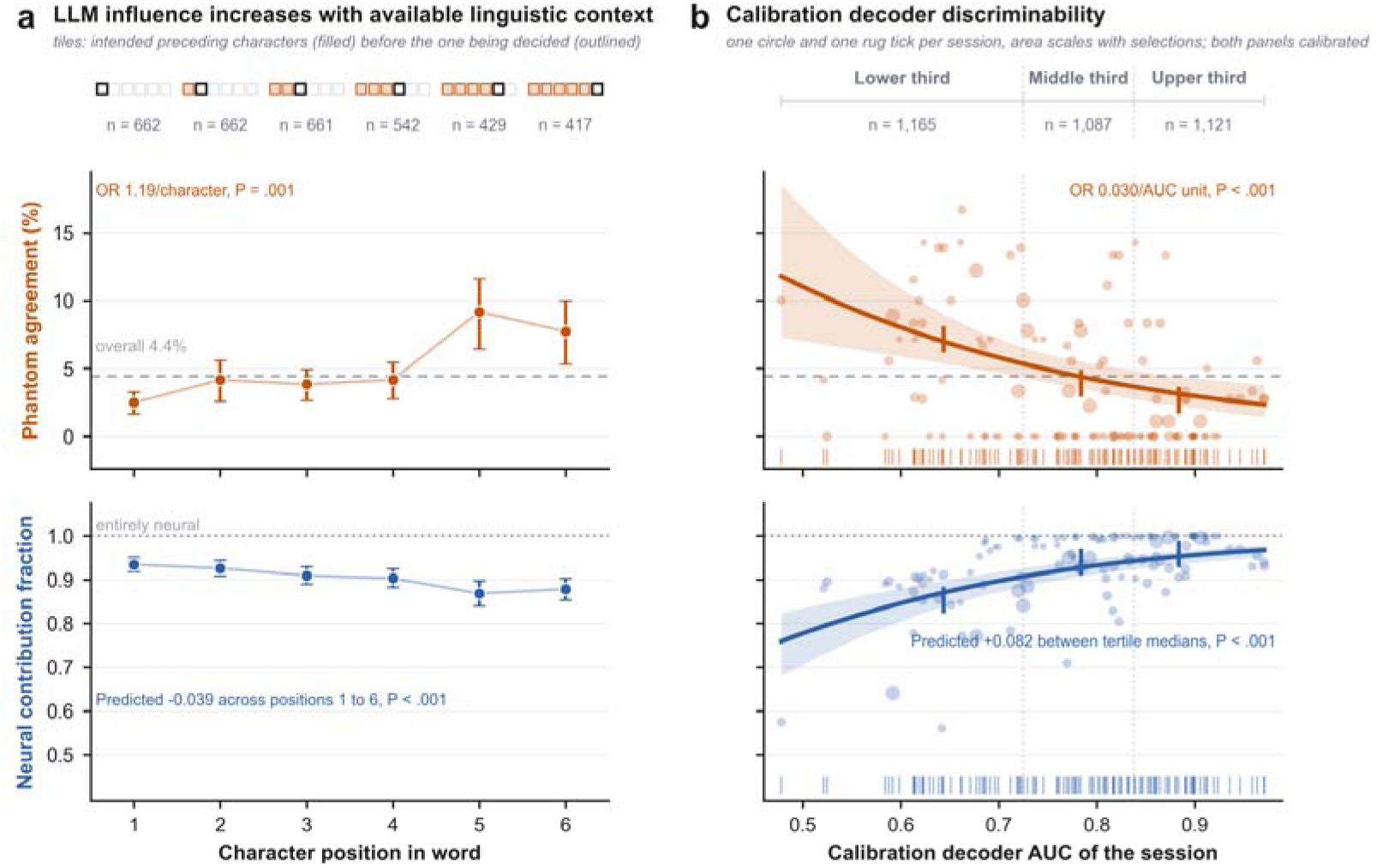
Attribution shifts toward the language model as context accumulates and as calibration decoder discriminability decreases. Both measures keep one color throughout: vermilion is phantom agreement, blue is the neural contribution fraction, and gray marks reference lines. The two panels share a y-axis range within each row. (a) The two measures by character position within the intended word. The tile strip above the panels shows what the prior had to condition on at each position, with the contributing selection count given beneath it; later positions occur only in longer words and so rest on fewer selections. The dashed gray line is the overall phantom agreement rate. (b) The same two measures against the calibration decoder AUC of the session, treated as the continuous quantity it is rather than as three bins; the fitted curve is the participant-clustered model reported in the Results (logistic for phantom agreement, a bounded fractional-logit fit for the neural contribution fraction that cannot predict outside [0, 1]) and the band is that model’s own 95% interval for the fitted mean. The small unlabeled ticks mark each tertile’s median AUC and point estimate, the value the Results quote, and the dotted verticals are the tertile boundaries. Error bars on the estimates are 95% confidence intervals from 2,000 participant-cluster bootstrap replicates. All estimates, curves, points, and annotations in both panels are on the calibrated-fusion basis of Figure 1 and the Results; raw, uncalibrated counterparts are the sensitivity comparisons in eTable 2 (position), eTable 3 (decoder quality), and eTable 5 (hypothesis tests). The neural contribution fraction’s annotation in each panel is the bounded fit’s own predicted change, across the plotted character positions in (a) and between the lower and upper tertiles’ median AUC in (b), not a per-unit slope from the separate linear mixed model reported as a sensitivity comparison in the Results and eTable 30. Annotated P values are each model’s own, unadjusted for multiplicity; Benjamini-Hochberg-adjusted values for the five-member secondary family are in the Results and eTable 30.

**Figure 3.**
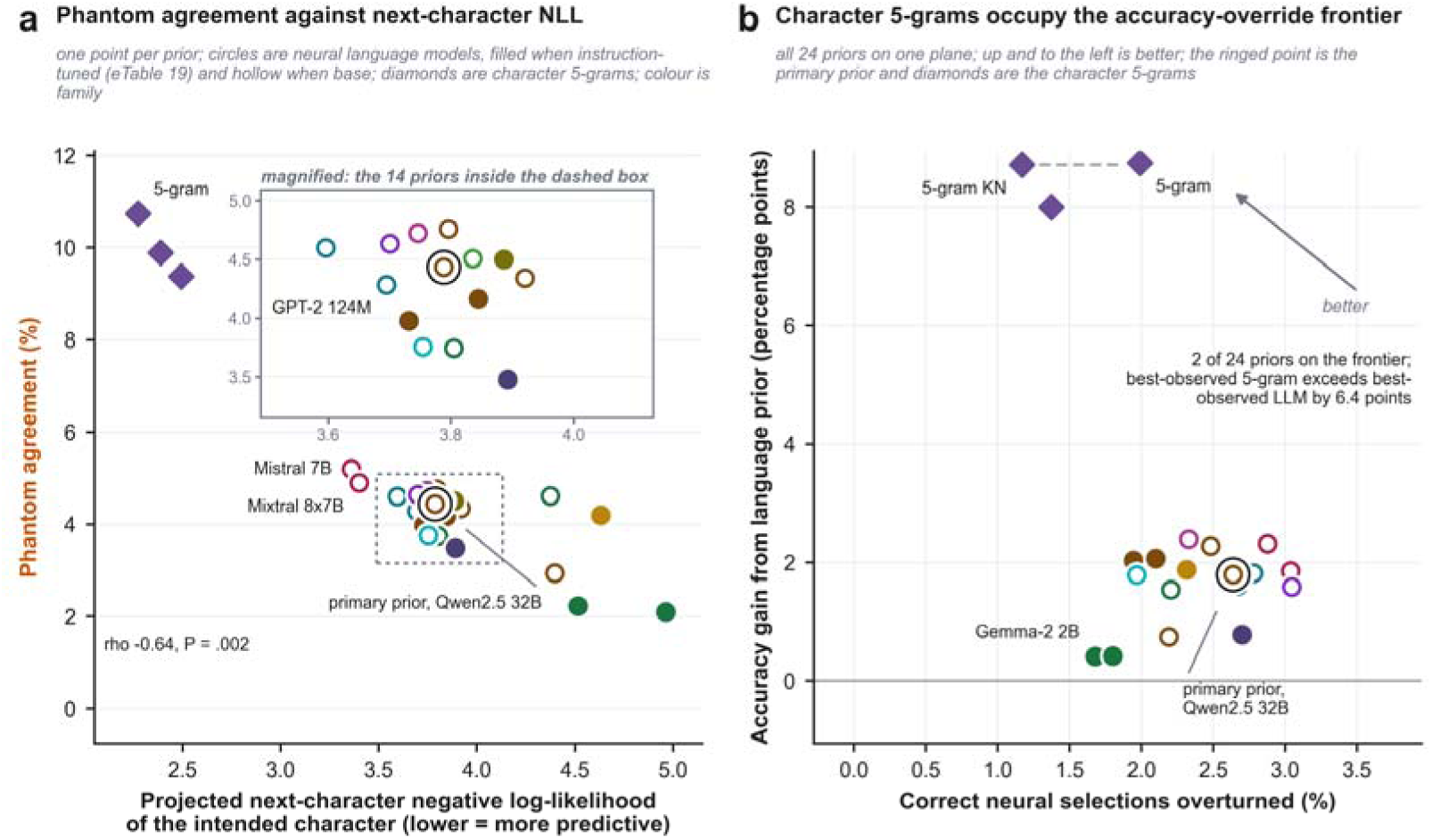
Next-character predictive quality is associated with phantom agreement, whereas no monotonic association with parameter count was detected, and character 5-grams alone occupy the frontier of accuracy gained against correct neural decisions overturned. One point per prior in both panels. The ringed point is the primary prior, Qwen2.5-32B. Both panels use the calibrated, held-out fusion basis of Figure 1 and Table 2; the raw basis is eTable 22’s sensitivity comparison. (a) The densest cluster sits in a dashed box, magnified in the inset for individual reading. Across the 21 neural language models, phantom agreement correlates with next-character NLL at rho = -0.64 (P = .002; eTable 25), a descriptive check, not the prespecified parameter-count equivalence test (eTable 18, eTable 23). Restricting the correlation to that class keeps predictive quality from being conflated with model class: the 5-grams predict characters natively, the unit the speller selects, whereas the neural language models are general-purpose checkpoints scored without task-specific adaptation. Across all 24 priors the correlation is stronger (rho = -0.76, P < .001), reported here as a descriptive comparison rather than in-panel because it spans both classes. Labeled: the primary prior, the best-NLL neural language model and 5-gram, and the largest and smallest neural language models by parameter count. The primary is labeled on the panel itself even inside the dashed box; the rest, in the inset when inside it. (b) What each prior buys against what it costs, defined on the panel’s own axes. Prior capture here is per-prior, from the calibrated ladder (Table 2); the ringed point’s prior capture is the primary-prior estimate reported in Results. The dashed step marks the benefit-harm frontier, the two points no prior beats on both axes: both are character 5-grams; the best of them, ngram5, exceeded the best neural model, DeepSeek-V2-Lite, by 6.4 percentage points on calibrated fused accuracy, a descriptive, not prespecified, comparison. Parameter counts for panel a’s largest/smallest labels are totals, including inactive experts of the four sparsely activated models (eTable 19).

**Figure 4.**
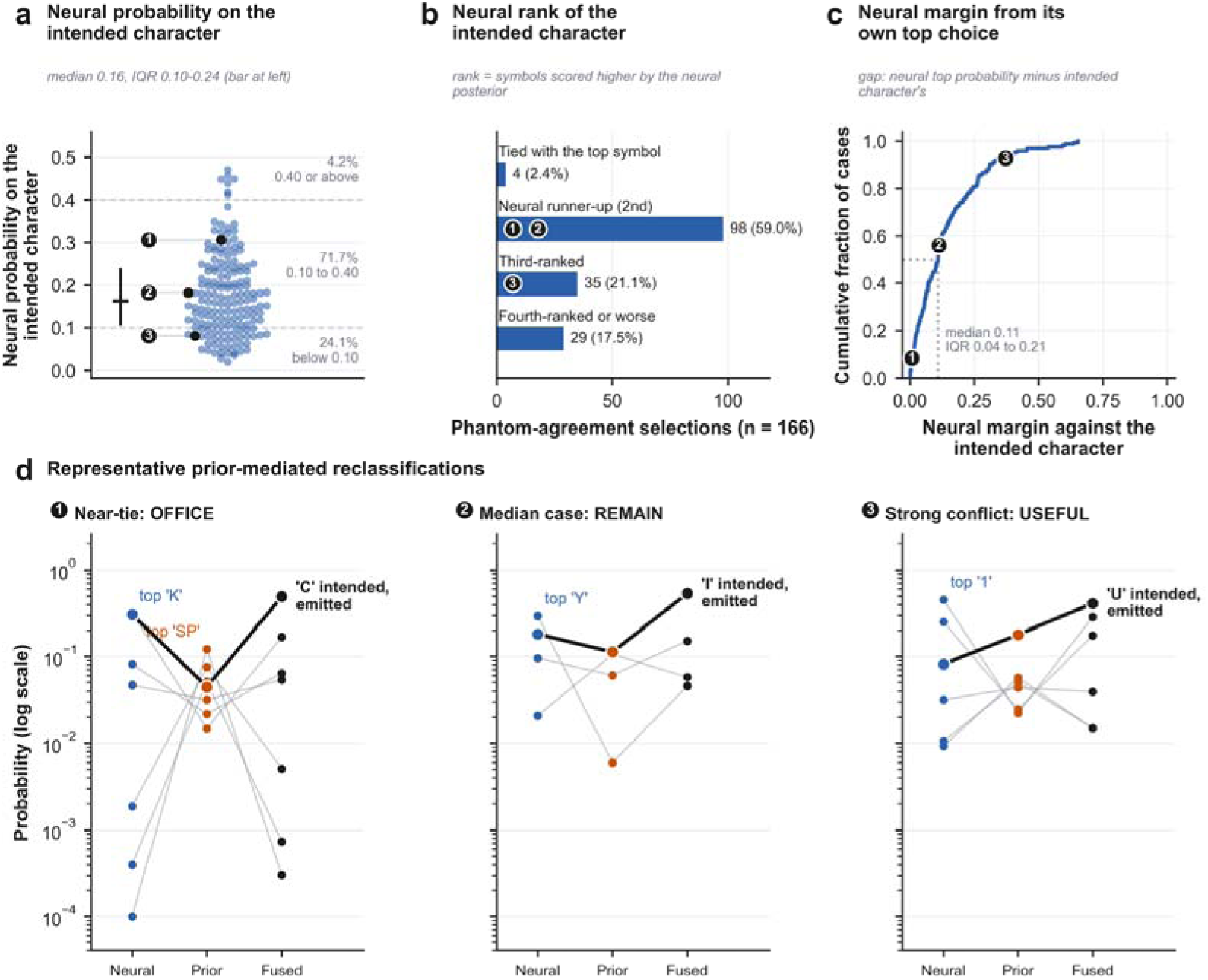
Neural support among prior-mediated reclassifications: how much evidence the intended character had in the 166 selections the calibrated fusion rule corrected. Every selection in which the fused posterior emitted the intended character and the neural posterior alone would not have; blue is neural evidence, vermilion the language-model prior, and charcoal the fused decision. (a) The neural probability on the intended character, one point per selection. (b) Where the intended character stood in the neural posterior’s own ranking; the 4 cases tied with its top symbol are shown separately, because a tie is resolved by a deterministic tie-break that can select against the intended character, and all four categories are shares of the full cohort (eTable 26). (c) The cumulative distribution of the neural margin. (d) Three of these selections in full, the ones nearest the 10th, 50th, and 90th percentiles of that margin among selections with at least four preceding characters, chosen by a rule fixed in advance. Numbered badges tie each card to its position in (a) to (c), and each source’s own top-ranked symbol is labeled above its column. The first, typing “OFFI” toward the intended “C” (OFFICE), is a near-tie the prior settled at a neural margin of 0.01 (8th percentile); the intended character was already the neural posterior’s own runner-up. The second, typing “REMA” toward the intended “I” (REMAIN), is the median case at a neural margin of 0.11 (56th percentile), the intended character again its own runner-up. The third, at the 93rd percentile of the cohort’s margins, is one in which the neural posterior favored a different symbol by a wide margin, and so is atypical of the rest of the cohort Probabilities in (d) are on a logarithmic scale and values below 10^−4^ are drawn at that floor. All panels are computed on the calibrated, held-out per-source fusion basis of Figure 1’s co-primary estimates and of Table 2, not the raw, uncalibrated basis eTable 11 reports as a disclosed sensitivity comparison.

#### Model influence also increased as calibration-decoder discriminability decreased

Across ascending tertiles of calibration AUC, phantom agreement was 7.2%, 3.9%, and 2.7%, and the neural contribution fraction was 0.853, 0.941, and 0.959 (Figure 2B; eTable 28). Phantom-agreement odds fell with calibration AUC (OR 0.030, 95% CI 0.005-0.204; *P*<.001; adjusted *P*<.001). The bounded model predicted a 0.082 increase in neural contribution between the outer-tertile median AUCs (0.056-0.109; *P*<.001; linear mixed-model slope +0.442 per AUC unit, adjusted *P*<.001; eTable 30), indicating a gradient rather than a threshold.

#### No statistically detectable difference in phantom agreement was observed between the prespecified digit-string and word strata

Calibrated phantom agreement was 6.6% versus 4.3% (OR 1.45, 95% CI 0.70-3.02; *P*=.32; adjusted *P*=.32; eTables 28 and 30).

### Association with parameter count

#### No monotonic association with parameter count was detected

Across 21 neural language models, calibrated phantom agreement ranged from 2.1% to 5.2% and was uncorrelated with log parameter count (Spearman rho -0.03; *P*=.91). The same was true for neural contribution fraction (rho -0.04; *P*=.86), prior capture (rho -0.08; *P*=.74), and accuracy gain (rho +0.18; *P*=.45) (Table 2; eTable 23; raw-basis figure in eFigure 4).

Qwen2.5-32B was used as the reference prior; its selection and subsequent expansion of the model ladder are documented in Supplementary Note S1.17. The full ladder comprised 21 neural language models from 11 architecture families, seven instruction-tuned and 14 base models.

Across the ladder, no attribution measure varied reliably with parameter count on either the raw or calibrated basis (eTables 9 and 23). An additional architecture search screened RWKV-7, xLSTM, StripedHyena, RetNet, and Gated DeltaNet; none passed the prespecified loadability check (eTable 10). Exact checkpoints, parameter counts, tuning status, and source organizations are in eTable 19.

Restricting the ladder to models below 10 billion parameters (11 of 21) showed no calibrated correlation for any measure (rho +0.01 to +0.31; *P*>=.36). A marginal raw-basis correlation for phantom agreement (rho +0.62; *P*=.044) did not persist after calibration (eTables 9 and 23).

Architecture-family permutation tests likewise found no measure with between-family variance above its null distribution on the calibrated basis (F-like statistic 0.92-2.01; *P*=.12-.53). However, predefined equivalence tests against a rho margin of +/-0.3 were inconclusive for every measure on both bases because all 90% CIs crossed the margin (eTables 18 and 31). These results show failure to detect a scale or family effect, not evidence of equivalence.

Character 5-grams produced the largest accuracy gains. Calibrated fused accuracy was 80.4% for the plain and Kneser-Ney 5-grams and 79.6% for the WikiText-103 5-gram, compared with a 71.6% uniform-prior baseline and 72.1-74.0% across neural language models (Table 2; raw fusion in eTable 22).

### Sensitivity analyses

Both co-primary findings persisted across prespecified sensitivity analyses. Raw, uncalibrated equal-weight fusion gave a neural contribution fraction of 0.925 (95% CI 0.907-0.943) and phantom agreement of 3.8% (3.0-4.5), both *P*<.001; calibration modestly increased the estimated prior share and phantom agreement (eTable 21). Across the fusion-exponent grid, calibrated phantom agreement ranged from 1.7% to 5.8% and raw phantom agreement from 1.4% to 5.7% (eFigure 1). In the highest calibration-quality tertile, phantom agreement remained 2.7% (1.5-3.7) and the neural contribution fraction 0.959 (n=1,121; eTable 28). Leave-one-study-out, stimulation-condition, and pooled-calibration exclusions produced the same qualitative pattern (eTable 32; eFigure 2; eTables 7-8 and 17).

A stratified context-permutation negative control paired each selection with a context from the same character-position, study, and context-length stratum. Among 2,710 selections with an eligible donor, the true context produced more phantom agreement and slightly lower neural contribution than 200 permuted context sets: 5.6% versus 3.3% (95% permutation interval 3.0-3.7) and 89.7% versus 90.1% (89.9-90.2). Both one-sided exceedance *P* values were <.005 (eTable 29), supporting context-specific linguistic influence rather than generic posterior sharpening.

Reduced numerical precision preserved the top-ranked symbol in 98.5% of contexts (Supplementary Note S1.3). All prespecified statistical models converged without fallback substitution.

## Discussion

In archived ALS P300-speller sessions, the language-model prior accounted for a participant-weighted mean 8.6% of posterior displacement across selections, with a strongly right-skewed distribution (median 2.9%). About 1 selection in 23 emitted the intended character although neural evidence alone would have selected another. Neither quantity is visible in accuracy or information-transfer rate. A system can therefore emit the intended character while its decision depends materially on the language prior.

The 8.6% estimate is specific to the Kullback-Leibler attribution index. Alternative formalisms yielded different numerical magnitudes, confirming that this is an operational index of posterior dependence rather than a unique decomposition of causal or human contribution (eTable 14).

Two gradients were consistent with the proposed interpretation. Model influence increased as more linguistic context accumulated within a word and as calibration-decoder discriminability decreased. Both position effects survived adjustment for participant, word, and session; the neural-contribution result also held when restricted to six-character words (eTables 20 and 28). The digit-string comparison was inconclusive.

We detected no monotonic association with parameter count and no architecture-family effect, but equivalence tests were inconclusive; these data therefore do not establish scale invariance. Predictive quality was more informative. Among 21 neural language models, lower next-character negative log-likelihood was associated with more phantom agreement (rho -0.64; *P*=.002; Figure 3). Character 5-grams also exceeded every neural model on calibrated fused accuracy (79.6-80.4% versus 72.1-74.0%). Under this character-projection interface, task-matched character models outperformed larger general-purpose checkpoints that had not been adapted to the task.

This analysis extends earlier mutual-information correction from completed strings to individual selections.^9^ It therefore identifies which correct characters were reclassified by the prior, not only how much prior information appeared in the final output. The results qualify claims that near-ceiling fused accuracy means the remaining bottleneck is solely neural decoding:^8^ accuracy can improve while some decisions become more dependent on the prior. The measures do not resolve the authorship, voice, or agency concerns raised by AAC users and clinicians.^11,12^ They quantify algorithmic dependence on the prior, not ownership, acceptance, voice preservation, ability to reject the output, or perceived agency.

Both measures can be calculated at run time from quantities already available to a fused speller. Devices could log the neural contribution fraction, flag selections that neural evidence alone would not have made, and expose those signals to users and clinical teams alongside accuracy. Such information may be most important when decoder quality is low, although calibration quality reflects hardware, recording conditions, and artefact burden rather than disease severity alone.

Several limitations apply. First, this was offline re-decoding, not observation of a person using an LLM-assisted speller. Participants could not see, accept, or correct the reconstructed characters, and no error-related-potential or adaptive feedback was available. An emitted-context sensitivity analysis allowed earlier errors to alter later context and reduced phantom agreement by 0.7 percentage points (eTables 16 and 27); intended-context analysis was therefore not uniformly an upper bound.

Second, the offline decoder was a reimplementation, with 71.8% accuracy versus 81.9% online and 69.6% agreement with archived selections. Weaker neural evidence increases the apparent contribution of the prior. Phantom agreement nevertheless remained 2.7% in the highest calibration-quality tertile, where offline selections reproduced 90.6% of archived outputs, and calibration AUC showed a graded association. The full-cohort magnitude may still overstate prior influence for a better-matched decoder.

Third, the task was copy-spelling of mostly six-character words rather than free composition, where longer and less constrained context could alter the prior’s influence.

Fourth, attribution depends partly on the fusion rule. Across the five-point neural-weighting grid, raw phantom agreement ranged from 1.4% to 5.7% and raw prior share from 3.7% to 56.6%; calibrated estimates varied non-monotonically from 1.7% to 5.8% and 8.4% to 89.9% (eFigure 1; eTable 6). The primary analysis used held-out, source-specific temperature calibration followed by equal weighting. Equal weight was chosen a priori rather than optimized against an external criterion, and one scalar temperature per source cannot correct miscalibration that varies by participant, context, or selection.

Fifth, the ladder extended to 46.7 billion parameters across 11 families but did not include frontier-scale or closed-weight systems. Equivalence tests could not exclude a moderate scale effect, and instruction tuning was confounded with model generation. The reference prior and ladder expansion were not fully prespecified; reference estimates should therefore be interpreted alongside the consistent results across all 21 neural language models (Supplementary Note S1.17). Parameter-count, architecture-family, and per-prior analyses are exploratory.

Sixth, the four source studies differed in stimulation design and electrode type, while age, sex, and ALSFRS-R were available only for subsets. Seventh, the digit-string stratum was small (168 selections versus 3,205 word selections), and short digit sequences may still be predictable; it is a weaker negative control than an unpredictable stimulus set, which the archive did not contain.

Eighth, next-symbol scoring depends on tokenizer behaviour. We used prefix-locked marginalization because re-tokenized continuation scoring changed the context tokenization in 33.9% of sampled context-candidate pairs (Supplementary Note S1.3b). All reported values use the prefix-locked scorer, but it has not been cross-validated against an independent tokenizer implementation, leaving residual tokenizer-specific uncertainty.

Future work should repeat the measurement online, relate it to user acceptance and correction behaviour, and extend it to free composition, frontier and closed-weight priors, and intracortical speech decoding, where language-model rescoring is common.

Influence from neural evidence and an assisting language prior can be measured for each selection. In this cohort it was neither negligible nor uniform: accuracy alone did not reveal how much the fused decision depended on the prior.

## Methods

### Study design and setting

This retrospective secondary analysis re-decoded archived online P300-speller sessions from people with amyotrophic lateral sclerosis (ALS) under offline language priors. None of the 25 priors evaluated here was used during the archived sessions. One archived condition used a separate online bigram prior to control dynamic stopping (eTable 8); excluding its 348 selections left both co-primary estimates materially unchanged on either fusion basis (eTable 17).

Reporting followed STROBE^14^ and the reporting checklist for language-model brain-computer interface studies.^15^

### Data source and cohort

Data came from BigP3BCI version 1.0.0, a public archive of online P300 studies containing raw electroencephalography, per-symbol stimulus channels, and feedback streams.^10^ We included source studies B, F, L, and N, each an ALS cohort with compatible calibration and online phases recorded on the shared 16-channel montage.

The unit of analysis was an online character selection. Eligibility required displayed feedback, recoverable intended and selected symbols, no artificial feedback overriding the classifier, at least two artefact-free flash epochs, and a contiguous preceding stimulation window. Three files were excluded: one target lay outside the 36-symbol grid and two lacked per-symbol flash channels. Because the task was copy-spelling, the intended character was recorded rather than inferred. The grid contained 26 letters, space, and digits 1-9. Flash membership was read from per-symbol channels because checkerboard stimulation can illuminate arbitrary symbol subsets. We verified the archive’s target-code mapping against readable copy-spelled words in all four studies (Supplementary Note S1.1).

### Neural posterior

A separate decoder was trained for each session using only its calibration phase. The 16 shared channels were filtered from 0.5 to 30 Hz with a fourth-order zero-phase Butterworth filter, epoched from -200 to 800 ms around flash onset, baseline-corrected on the prestimulus interval, and rejected if absolute amplitude exceeded 150 microvolts. Epochs were downsampled to approximately 20 samples per channel, standardized, and classified with an L2-regularized logistic model using balanced class weights (MNE-Python^16^). Calibration AUC was estimated by stratified grouped cross-validation held out by calibration file. Two sessions without calibration used pooled calibration files from the same participant.

For each online selection, flash-level log odds were summed onto every illuminated symbol and normalized across the 36-symbol alphabet: posterior proportional to exp(M transpose times s), where M is the flash-by-symbol membership matrix and s is the vector of flash log odds. The classifier receives EEG features only, never symbol identity, and therefore does not encode a natural letter- or digit-frequency prior.

Before scaling, both sources were overconfident: top-label expected calibration error was 16.7% for the neural posterior and 34.7% for the language-model prior. Held-out, source-specific temperature calibration reduced these values to 4.8% and 4.1%, with parallel improvements in negative log-likelihood and Brier score (eTable 24). These diagnostics do not establish the optimal relative fusion weight, which was examined separately with a fusion-exponent sensitivity analysis. Because the offline posterior reimplemented rather than reproduced the archived decoder, we measured its fidelity directly: its argmax matched the archived online selection in 69.6% of trials, compared with 2.8% by chance, and fidelity tracked calibration quality (eTable 3). The highest-fidelity tertile was therefore a key sensitivity analysis (Supplementary Note S1.5).

### Language-model priors

Priors were defined over the same 36-symbol alphabet and conditioned on the intended within-word prefix, the primary oracle-context specification. An emitted-context sensitivity analysis instead propagated the fused system’s earlier outputs within each word (Supplementary Note S1.14; eTable 16).

The ladder comprised 25 priors: a uniform null, three character 5-grams trained on the Brown corpus or WikiText-103,^17,18^ and 21 causal language models from 124 million to 46.7 billion parameters across 11 architecture families.^19–32^ Qwen2.5-32B was the reference prior for both co-primary outcomes. Its selection and the subsequent ladder expansion are documented in

Supplementary Note S1.17; results across all 21 neural language models are reported alongside the reference estimate. eTable 19 lists exact checkpoints, parameter counts, and tuning status. Small models were scored locally and larger models on an academic cluster; a reference model produced numerically equivalent priors in both environments (Supplementary Note S1.3).

### Next-symbol scoring

Models were used for likelihood scoring only; no sampling parameters or random seed were involved, and each distinct context was scored once. Because byte-pair tokens do not align with characters, the context was tokenized once and passed through the model once. The next-token distribution was then projected onto the 36 grid symbols according to each vocabulary token’s first character. Contexts were lower-cased, predicted characters were folded back to the grid alphabet, and reduced precision was used where required; its effect was quantified (Supplementary Note S1.3).

We rejected a comparator that separately re-tokenized each context-plus-candidate string. In a tokenizer-only diagnostic using 200 real contexts and all 36 candidate symbols, appending the candidate changed the context tokenization in 33.9% of 7,200 pairs. Prefix-locked marginalization removes that artefact by construction, and all reported prior- and fusion-dependent estimates use this scorer (Supplementary Note S1.3b).

### Fusion and posterior calibration

Neural and language-model distributions were combined by the standard Bayesian speller product, with the neural term raised to fusion exponent beta=1 in the primary analysis.

Each source was calibrated before primary fusion. A separate scalar temperature minimized mean negative log-likelihood of the intended symbol in five participant-grouped folds; every held-out participant was scored using temperatures fitted on the other four folds. Mean fitted temperatures were 3.12 for the neural posterior and 2.40 for the language-model posterior, indicating overconfidence in both sources (Supplementary Note S1.10b; eTable 21). Calibrated distributions were then fused at beta=1. An unfitted diagnostic grid applied a shared temperature T in {0.5, 1.0, 2.0} to both sources; raw, uncalibrated beta=1 fusion was retained as a labelled sensitivity comparison (Supplementary Note S1.10; eTable 13). Unless noted otherwise, secondary and sensitivity analyses used the held-out calibrated basis.

### Outcomes

Two co-primary constructs quantified dependence on the prior relative to neural evidence. With n the neural posterior, p the language prior, and f the fused posterior over 36 symbols, the neural contribution fraction was NCF = D(f || p) / [D(f || p) + D(f || n)], where D is Kullback-Leibler divergence. NCF lies in [0,1]: it equals 1 when the prior is uniform and f equals n, and 0 when neural evidence is uninformative and f equals p. It is undefined only if both sources are uniform, which did not occur. Phantom agreement indicated that the fused posterior emitted the intended symbol while the neural posterior alone would not have.

Secondary outcomes were prior capture (the converse reclassification), neural override (a correct fused emission selected by the neural posterior but not by the prior), and the change in accuracy after adding the prior. Phantom agreement and prior capture are mutually exclusive. Prespecified moderators were character position, calibration-decoder quality, target type, stimulation condition, and source study.

### Statistical analysis

Each selection contributed once to the reference-prior co-primary analyses. The Bonferroni-split alpha was 0.025; the secondary family was tested only after at least one co-primary endpoint met its threshold. Neural contribution fraction was tested one-sided against 1 and phantom agreement one-sided against 0 in the prespecified directions.

Point estimates were participant-weighted: each measure was calculated within participant and then averaged across participants. Confidence intervals and one-sided *P* values came from 2,000 deterministic participant-cluster bootstrap replicates that resampled participants with replacement.

Fractional-logit generalized estimating equations, constrained to [0,1] and summarized as predicted-value contrasts, provided the primary moderator estimates for neural contribution fraction. A linear mixed model with participant random intercept was retained as a sensitivity analysis and supplied adjusted *P* values for continuous multiplicity-family members. Binary outcomes used logistic generalized estimating equations and are reported as odds ratios with 95% CIs. Both models used exchangeable working correlation clustered by participant. The five-member secondary family used Benjamini-Hochberg adjustment^38^ at 0.05; a cluster-robust fallback was prespecified for non-convergence.

### Robustness and sensitivity analyses

Prespecified sensitivity analyses varied the fusion exponent from 0.25 to 4, restricted the cohort to the highest calibration-quality tertile, excluded pooled-calibration sessions, and removed each source study in turn.

Position effects were also refit with participant, word, and session represented as crossed effects. Neural contribution fraction was separately restricted to six-character words, the only length spanning all six positions. Phantom agreement used a Bayesian mixed-effects binomial model for the crossed analysis; its credible interval was reported separately from the Benjamini-Hochberg family (Supplementary Note S1.6b; eTable 20).

Additional robustness analyses included a six-scenario construct-validity simulation, two non-KL attribution formalisms, a context-permutation negative control, source-specific calibration diagnostics, architecture-family permutation testing, and equivalence tests for parameter-count associations (Supplementary Notes S1.9-S1.13 and S1.16; eTables 12, 14, 15, 24, and 31). These analyses tested whether the qualitative conclusion depended on the KL formulation, generic posterior sharpening, source miscalibration, architecture family, or the sampled parameter range.

## Ethics

BigP3BCI is an open-access, de-identified archive released under a Creative Commons Attribution 4.0 licence. The source P300 studies were approved by the institutional review boards of Duke University, Duke University Health System, and East Tennessee State University, and participants provided informed consent directly or through a legally authorized representative. The Beth Israel Deaconess Medical Center Human Research Protection Program determined that this secondary analysis of de-identified records constituted non-human-subjects research and required no additional institutional review board approval or consent. The study followed the Declaration of Helsinki.

## Data availability

BigP3BCI version 1.0.0 is publicly available from PhysioNet (doi:10.13026/0byy-ry86).

## Code availability

Analysis code is publicly available at https://github.com/BRIDGE-GenAI-Lab/BCI_Authorships; a permanent archive will be created on Zenodo/OSF upon publication.

## Acknowledgments

This work received no external funding.

## Generative-AI disclosure

ChatGPT (OpenAI, San Francisco, CA, USA) was used for language editing only. It was not used for study design, statistical analysis, interpretation, or generation of any reported estimate. The authors verified all content and take full responsibility for the manuscript.

## Author contributions

Contributions are described using the CRediT taxonomy. A.G.: conceptualization, methodology, software, formal analysis, data curation, validation, visualization, and writing of the original draft. M.O.: formal analysis and review and editing of the manuscript. E.J.: validation and review and editing of the manuscript. Y.A.: software, data curation, formal analysis, validation, and review and editing of the manuscript. O.D.: conceptualization, methodology, investigation, formal analysis, clinical interpretation, supervision, writing of the original draft, and review and editing of the manuscript. J.K.: supervision and review and editing of the manuscript. M.A.: resources and review and editing of the manuscript. O.B.: validation and review and editing of the manuscript. Y.B.: investigation and review and editing of the manuscript. E.K.: conceptualization, methodology, supervision, resources, and review and editing of the manuscript. All authors critically reviewed the manuscript and approved the final version submitted for publication. A.G. (corresponding author) had full access to all data in the study and takes responsibility for the integrity of the data and the accuracy of the analysis.

## References

1. Farwell L, Donchin E. Talking off the top of your head: toward a mental prosthesis utilizing event-related brain potentials. Electroencephalogr Clin Neurophysiol. 1988;70(6):510–523. doi:10.1016/0013-4694(88)90149-6

2. Vansteensel MJ, Pels EG, Bleichner MG, Branco MP, Denison T, Freudenburg ZV, et al. Fully Implanted Brain–Computer Interface in a Locked-In Patient with ALS. N Engl J Med. 2016;375(21):2060–2066. doi:10.1056/NEJMoa1608085

3. Speier W, Arnold C, Lu J, Deshpande A, Pouratian N. Integrating Language Information With a Hidden Markov Model to Improve Communication Rate in the P300 Speller. IEEE Trans Neural Syst Rehabil Eng. 2014;22(3):678–684. doi:10.1109/tnsre.2014.2300091

4. Speier W, Arnold CW, Deshpande A, Knall J, Pouratian N. Incorporating advanced language models into the P300 speller using particle filtering. J Neural Eng. 2015;12(4):046018. doi:10.1088/1741-2560/12/4/046018

5. Lebedev MA, Makarova AV, Kleeva DF, Maysuradze AI. Connecting a P300 speller to a large language model. bioRxiv preprint. doi:10.1101/2025.11.06.686984

6. Hong J, Wang W, Najafizadeh L. ChatBCI, a P300 speller BCI with context-driven word prediction leveraging large language models, from concept to evaluation. Sci Rep. 2026;16(1). doi:10.1038/s41598-025-25660-7

7. Paplavsky N, Lebedev M. Postprocessing of P300 Speller Output with a Large Language Model. bioRxiv preprint. doi:10.64898/2026.06.24.734268

8. Parthasarathy N, Soetedjo J, Panchavati S, Parthasarathy N, Lee D, Arnold C, et al. Near-Optimal P300 Speller Performance Using Large Language Models: A Multi-Model Analysis with Performance Bounds. bioRxiv preprint. doi:10.1101/2025.10.28.685216

9. Speier W, Arnold C, Pouratian N. Evaluating True BCI Communication Rate through Mutual Information and Language Models. PLoS One. 2013;8(10):e78432. doi:10.1371/journal.pone.0078432

10. Mainsah B, Fleeting C, Balmat T, Sellers E, Collins L. bigP3BCI: An Open, Diverse and Machine Learning Ready P300-based Brain-Computer Interface Dataset. PhysioNet; 2025. doi:10.13026/0byy-ry86

11. Valencia S, Cave R, Kallarackal K, Seaver K, Terry M, Kane SK. “The less I type, the better”: How AI Language Models can Enhance or Impede Communication for AAC Users. In: Proceedings of the 2023 CHI Conference on Human Factors in Computing Systems. 2023:1–14. doi:10.1145/3544548.3581560

12. Weinberg TM, Gonzalez Penuela RE, Valencia S, Roumen T. I, Robot? Exploring Ultra-Personalized AI-Powered AAC; an Autoethnographic Account. In: Proceedings of the 2026 CHI Conference on Human Factors in Computing Systems. 2026:1–16. doi:10.1145/3772318.3790310

13. Xie Y, Qi T, Yi J, Yang X, Whalen R, Huang J, et al. Measuring Human Contribution in AI-Assisted Content Generation. arXiv preprint arXiv:2408.14792.

14. von Elm E, Altman DG, Egger M, Pocock SJ, Gøtzsche PC, Vandenbroucke JP. The Strengthening the Reporting of Observational Studies in Epidemiology (STROBE) statement: guidelines for reporting observational studies. Lancet. 2007;370(9596):1453–1457. doi:10.1016/S0140-6736(07)61602-X

15. Gorenshtein A, Omar M, Barash Y, Nadkarni GN, Klang E. Large language models integrated into brain–computer interfaces for communication and control: a systematic review. Biomed Phys Eng Express. 2026;12(3):035077. doi:10.1088/2057-1976/ae737b

16. Gramfort A. MEG and EEG data analysis with MNE-Python. Front Neurosci. 2013;7. doi:10.3389/fnins.2013.00267

17. Kneser R, Ney H. Improved backing-off for M-gram language modeling. 1995 International Conference on Acoustics, Speech, and Signal Processing. 1995;1:181–184. doi:10.1109/ICASSP.1995.479394

18. Merity S, Xiong C, Bradbury J, Socher R. Pointer Sentinel Mixture Models. arXiv preprint arXiv:1609.07843.

19. Yang A, Yang B, Zhang B, Hui B, Zheng B, Yu B, et al. Qwen2.5 Technical Report. arXiv preprint arXiv:2412.15115.

20. Qwen Team. Qwen3.5: Towards Native Multimodal Agents. Published February 2026. https://qwen.ai/blog?id=qwen3.5

21. Qwen Team. Qwen3.6-35B-A3B: Agentic Coding Power, Now Open to All. Published April 2026. https://qwen.ai/blog?id=qwen3.6-35b-a3b

22. Grattafiori A, Dubey A, Jauhri A, Pandey A, Kadian A, Al-Dahle A, et al. The Llama 3 Herd of Models. arXiv preprint arXiv:2407.21783.

23. Riviere M, Pathak S, Sessa PG, Hardin C, Bhupatiraju S, Hussenot L, et al. Gemma 2: Improving Open Language Models at a Practical Size. arXiv preprint arXiv:2408.00118.

24. Abd SE, Aggarwal V, Algayres R, Andreev A, Bachem O, Ballantyne I, et al. Gemma 4 Technical Report. arXiv preprint arXiv:2607.02770.

25. Jiang AQ, Sablayrolles A, Mensch A, Bamford C, Chaplot DS, de las Casas D, et al. Mistral 7B. arXiv preprint arXiv:2310.06825.

26. Jiang AQ, Sablayrolles A, Roux A, Mensch A, Savary B, Bamford C, et al. Mixtral of Experts. arXiv preprint arXiv:2401.04088.

27. Liu A, Feng B, Wang B, Wang B, Liu B, Zhao C, et al. DeepSeek-V2: A Strong, Economical, and Efficient Mixture-of-Experts Language Model. arXiv preprint arXiv:2405.04434.

28. Walsh P, Soldaini L, Groeneveld D, Lo K, Arora S, Bhagia A, et al. 2 OLMo 2 Furious. arXiv preprint arXiv:2501.00656.

29. Agarwal S, Ahmad L, Ai J, Altman S, Applebaum A, Arbus E, et al. gpt-oss-120b & gpt-oss-20b Model Card. arXiv preprint arXiv:2508.10925.

30. Biderman S, Schoelkopf H, Anthony QG, Bradley H, O’Brien K, Hallahan E, et al. Pythia: A Suite for Analyzing Large Language Models Across Training and Scaling. Proceedings of Machine Learning Research. 2023;202:2397–2430. doi:10.48550/arXiv.2304.01373

31. Gu A, Dao T. Mamba: Linear-Time Sequence Modeling with Selective State Spaces. arXiv preprint arXiv:2312.00752.

32. De S, Smith SL, Fernando A, Botev A, Cristian-Muraru G, Gu A, et al. Griffin: Mixing Gated Linear Recurrences with Local Attention for Efficient Language Models. arXiv preprint arXiv:2402.19427.

33. Peng B, Zhang R, Goldstein D, Alcaide E, Du X, Hou H, et al. RWKV-7 “Goose” with Expressive Dynamic State Evolution. arXiv preprint arXiv:2503.14456.

34. Beck M, Poppel K, Lippe P, Kurle R, Blies PM, Klambauer G, et al. xLSTM 7B: A Recurrent LLM for Fast and Efficient Inference. arXiv preprint arXiv:2503.13427.

35. Poli M, Massaroli S, Nguyen E, Fu DY, Dao T, Baccus S, et al. Hyena Hierarchy: Towards Larger Convolutional Language Models. arXiv preprint arXiv:2302.10866.

36. Sun Y, Dong L, Huang S, Ma S, Xia Y, Xue J, et al. Retentive Network: A Successor to Transformer for Large Language Models. arXiv preprint arXiv:2307.08621.

37. Yang S, Kautz J, Hatamizadeh A. Gated Delta Networks: Improving Mamba2 with Delta Rule. arXiv preprint arXiv:2412.06464.

38. Benjamini Y, Hochberg Y. Controlling the False Discovery Rate: A Practical and Powerful Approach to Multiple Testing. J R Stat Soc Series B Stat Methodol. 1995;57(1):289–300. doi:10.1111/j.2517-6161.1995.tb02031.x

