## appendix for "Quantifying Large Language Model Influence in Brain Computer Interface Communication for Amyotrophic Lateral Sclerosis"

This Supplementary Information accompanies the main manuscript. It contains the full attribution formalism, the decoder and language-model scoring detail, the validation evidence that supports the reconstruction, the per-stratum result tables, and the sensitivity analyses summarized in the main text.

**Contents**

- S1 Supplementary Validation and Implementation Notes
- S2 Supplementary Tables (eTable 1 to eTable 32)
- S3 Supplementary Figures (eFigure 1 to eFigure 4)
- S4 Code and Data Availability

### S1 Supplementary Validation and Implementation Notes

#### S1.1 Grid alphabet and flash membership

Each recording exposes one channel per symbol, named by symbol, row and column; these are the authoritative record of which symbols were illuminated on each flash (Methods). In the analysed recordings, checkerboard flashes illuminated 3 to 5 symbols, most often 4, and the intended symbol appeared in 11.1% of flashes, which matches the 4-of-36 expectation.

The archive's target-code-to-symbol mapping (Methods) was verified against the data: concatenating the intended symbols of consecutive selections reproduced readable copy-spelling words in all four source studies, including THE, LAZY and DOG in Study B, ENOUGH and WINDOW in Study F, PLENTY, THREAT, KNIGHT and ADVICE in Study L, and VALLEY, PLACES, SINGLE and SENIOR in Study N.

#### S1.2 Decoder and neural posterior

Feature standardization, applied before classification (Methods), is not merely conventional: an unscaled logistic fit on the raw epoch features is numerically unstable.

The calibration-file holdout used for cross-validation (Methods) ensured that no epoch was ever scored by a model fitted on its own recording.

#### S1.3 Language-model priors and the choice of next-symbol scorer

Two next-symbol scorers were compared before selecting the prefix-locked marginalization used throughout (Methods): batched continuation scoring, which re-tokenizes the context separately for each of the 36 candidate symbols, and prefix-locked marginalization, which tokenizes the context once and reads all 36 candidate masses off a single forward pass through a static first-character projection built once per model. There is no per-candidate step of any kind in the production scorer: one tokenization, one forward pass, one matrix product.

The two methods disagree substantially. Running both over all 951 distinct copy-spelling contexts, for the three models scored under both scorers (GPT-2, GPT-2 large and Qwen2.5-1.5B), the top-ranked symbol agreed in only 7.6%, 10.8% and 13.4% of contexts, and the mean absolute difference across the alphabet was 0.044, 0.043 and 0.038, respectively. The remaining models, including the primary prior, were rescored under the corrected method rather than run under both, so this context-level comparison is reported for those three models and is not asserted for the ladder as a whole. The space-symbol mass shift disclosed in Methods arises because a model asked for the next token after a truncated word frequently predicts a word boundary.

An earlier revision read this disagreement as evidence against the marginalization and used batched continuation scoring throughout; that reading did not survive the tokenizer diagnostic reported in S1.3b and was reversed (Methods). Continuation scoring is retained in the code only as a labelled comparator, never in the production path. Either method is affordable here because copy-spelling contexts repeat: 3,373 selections required only 951 distinct contexts, each scored once per model.

Priors for the four smallest models (GPT-2, GPT-2 large, Qwen2.5-1.5B, and Qwen2.5-3B, up to 3 billion parameters) were scored on a local workstation. The remaining 17 neural models, from 2 to 46.7 billion parameters (Qwen2.5-14B, Qwen3.5-27B, Qwen2.5-32B, Qwen3.6-35B-A3B, Llama-3.1-8B, Gemma-2-2B, Gemma-2-9B, Gemma-2-27B, Gemma-4-12B, Mistral-7B, Mixtral-8x7B, DeepSeek-V2-Lite, OLMo-2-7B, GPT-OSS-20B, Pythia-12B, Mamba-2.8B, and RecurrentGemma-2B), were scored on an academic computing cluster using 1 to 3 NVIDIA L40S graphics processors depending on model size, with Mixtral-8x7B requiring 3, GPT-OSS-20B requiring 2, and most others requiring 1, using the same code and the same context set. Gemma-2-2B, Mamba-2.8B, and RecurrentGemma-2B were scored on the cluster with their round of models despite being small enough to qualify for local scoring by parameter count alone. Because that introduces a second software and hardware environment, a reference model was re-scored on the cluster and compared against its local priors over all 951 contexts: the top-ranked symbol agreed in 100% of contexts and the mean absolute difference across the alphabet was 1.4 times ten to the power minus 7, which is float32 rounding. The two environments are therefore interchangeable.

Reduced precision was used where memory required it. Its effect was quantified directly by scoring the 1.5-billion-parameter model in both precisions over all 951 contexts: the top-ranked symbol agreed in 98.5% of contexts, the mean absolute difference across the alphabet was 0.00045, and the mean absolute divergence was 0.0007 bits.

#### S1.3b Prefix-tokenization stability of batched continuation scoring

Batched continuation scoring (S1.3) re-tokenizes the complete string "context plus candidate symbol" separately for each of the 36 candidates and normalizes the 36 resulting total log-probabilities against one another. Treating that normalization as scoring a single next-symbol distribution conditioned on a fixed context implicitly assumes that the context's own tokenization does not change when a candidate symbol is appended to it, i.e. that the context's own tokenization is a strict prefix of the context-plus-candidate's tokenization for every candidate. Byte-pair tokenizers are not guaranteed to satisfy this: because tokenization is re-derived from the full string rather than extended token-by-token, appending a character can redraw an earlier token boundary.

This was tested directly rather than assumed, with a CPU-only, tokenizer-only diagnostic requiring no model forward pass and no GPU. A random sample of 200 of the 951 distinct real context-prefix values (fixed seed 20260805) was tokenized both alone and with each of the 36 grid symbols appended, lower-cased exactly as the primary prior presents context (S1.3), using the identical tokenizer object the primary prior loads (Qwen2.5-32B's own tokenizer, matching how the primary prior loads it). Of the 7,200 context-candidate pairs checked, 66.1% left the context's tokenization unchanged and 33.9% did not: for example, "remot" tokenizes as two tokens, but "remot" + "e" tokenizes as one whole-word token unrelated to either, and "a" tokenizes as one token that is not a prefix of "ab"'s single merged token.

This was a real and substantial property of the batched re-tokenization scorer used in an earlier revision of this analysis, not a rare edge case: for roughly one in three candidate symbols scored, the log-probability mass attributed to the shared context was not in fact a constant that cancels when the 36 totals are normalized, so a symbol whose appended character happened to trigger a cheaper (or costlier) re-tokenization of the context could gain or lose posterior mass for reasons unrelated to how linguistically plausible that symbol is. This revision replaces that scorer with prefix-locked marginalization, which removes the artefact by construction rather than by correction, not by correcting for it after the fact (Methods). Every prior in the ladder was rescored under this implementation, and every estimate reported in this manuscript reflects it (Methods).

#### S1.4 Attribution formalism

**Neural contribution fraction.** Displacement (Methods) is measured in bits. The bound, equality cases, and the one undefined case are given in Methods; none of the three was clipped or handled as a special case in the implementation; each follows directly from the formula.

**Categorical measures.** With k denoting the intended symbol:

| Measure | Definition |
| --- | --- |
| Phantom agreement | argmax f equals k and argmax n does not equal k |
| Prior capture | argmax n equals k and argmax f does not equal k |
| Neural override | argmax f equals k and argmax n equals k and argmax p does not equal k |

Mutual exclusivity of phantom agreement and prior capture (Methods) was asserted directly on the analysis frame rather than assumed.

**Fusion.** Follows the fixed-exponent Bayesian product (beta = 1 primary; 0.25 to 4 sensitivity grid) described in Methods.

#### S1.5 Reconstruction fidelity

The offline posterior is a reimplementation and does not exactly reproduce the archive's online decoder; Methods reports the argmax-match fidelity, its chance baseline, and its association with calibration quality (eTable 3). Fidelity was prespecified as the stratifying variable for the highest-fidelity-tertile sensitivity analysis because weaker neural evidence inflates the apparent contribution of the prior, so uncontrolled reconstruction quality could otherwise be mistaken for a genuine attribution effect.

#### S1.6 Statistical inference

Model specification, the co-primary testing plan, and the bootstrap procedure are described in Methods (Statistical Analysis). Two points here are implementation detail beyond that description.

Binary outcomes were represented as integers before modelling: a boolean outcome is expanded by the formula layer into a two-level categorical whose first level is false, which silently models the complement and inverts every odds ratio. A regression test guards this behaviour in the analysis code.

All models converged; no prespecified analysis required the planned cluster-robust substitution.

Separately, the categorical argmax measures defined in S1.4 (neural argmax, prior argmax, fused argmax) resolve exact ties toward the lower symbol index, following numpy.argmax's documented first-occurrence behaviour rather than any randomized tie-break, so this is deterministic and unaffected by the bootstrap seed. Exact ties are expected to be exceedingly rare given the continuous-valued neural, prior, and fused posteriors.

#### S1.6b Word- and session-adjusted position-in-word effect

The eTable 2 model carries only a participant random intercept, so it cannot rule out two further sources of clustering: the same word recurring across the corpus, and clustering within a recording session. statsmodels.MixedLM's groups= argument supports only one nested hierarchy, so representing this crossed structure (the same word appearing across many different participants' data, rather than nested inside one) used a variance-components mechanism instead: every observation was assigned to a single dummy top-level group, and participant, word identity, and recording session were each fitted as an independent, fully crossed variance component. Session identifier alone is not a valid grouping key for this purpose: its values are recycled per participant (every participant's own first recorded session is labelled "SE001", the second "SE002", and so on, across only 9 distinct raw labels), so a composite participant-by-session key was built first to give each of the archive's 115 recording sessions its own identity.

The default optimizer did not reach a reported convergence for this specification. The lbfgs optimizer did (maxiter=500), and three further optimizers (conjugate gradient, Powell, Nelder-Mead) converged independently to the same position coefficient reported in Methods (-0.0068) to four decimal places, so that estimate rests on four independent fits rather than one unverified result. Had no optimizer converged, the planned cluster-robust substitution (Methods) would have applied; it was not needed here. A second, independent check refit the unadjusted eTable 2 model (unchanged, participant random intercept only, the same fitting call used throughout) restricted to the 2,502 selections from six-character words, the corpus's only word length spanning all six positions, removing the possibility that the gradient reflects word length rather than position itself. Both results are reported in eTable 20 together with the crossed model's three variance-component estimates, so the size of the word and session effects it adjusts for is visible alongside the position estimate itself.

Phantom agreement, the binary co-primary outcome, carries its own position-in-word gradient and was adjusted for the same crossed structure separately. The logistic generalized estimating equations used elsewhere for this outcome admit only one clustering variable, so word identity and session cannot be adjusted for alongside participant there. The same variance-components approach can be applied with a binomial family, but it estimates a variational (mean-field Gaussian) approximation to the posterior rather than solving estimating equations. The interval this produces is therefore a 95% credible interval from that approximate posterior, not a sandwich-based confidence interval, and no P value is defined for it, which is why this fit is excluded from the Benjamini-Hochberg-adjusted secondary family (eTable 5, eTable 30) and carries no adjusted P value.

statsmodels jitters the variational fit's starting posterior standard deviations off the global NumPy random state, which would write a different coefficient into the tracked digest on every run; the unjittered centre of that draw is passed as an explicit start instead, removing the dependence on global random state and reaching the same optimum. No six-character-words-only refit is reported for phantom agreement, so word length is addressed for the neural contribution fraction only. Results are in eTable 20 (raw fusion) and eTable 28 (calibrated fusion).

#### S1.7 Implementation notes

The continuous attribution measure is the Kullback-Leibler displacement share described in S1.4 rather than a raw log-evidence ratio: the ratio form is unbounded and would need clipping, while the displacement share is bounded by construction and has the correct limiting behaviour at both endpoints.

The neural language model uses first-token marginalization throughout (S1.3); an alternative, batched continuation scoring, was evaluated and set aside once the tokenizer diagnostic in S1.3b showed it rests on a prefix-invariance assumption that fails for a third of sampled pairs.

The emitted-context sensitivity analysis (Supplementary Note S1.14, eTable 16) is reported for the primary prior only, because emitted context depends on the fusion rule under test and would make the context differ between arms of the prior ladder.

#### S1.8 Software

Local analyses used Python 3.9.6 with NumPy 2.0.2, SciPy 1.13.1, scikit-learn 1.6.1, statsmodels 0.14.6, pandas 2.3.3, MNE 1.8.0, transformers 4.57.6, PyTorch 2.8.0, and NLTK 3.9.2, with language-model scoring on Apple Metal hardware. Cluster scoring used Python 3.11 with transformers 5.14.1, PyTorch 2.6.0 and CUDA 12.4 on NVIDIA L40S hardware. Equivalence of the two environments is reported in S1.3.

#### S1.9 NCF construct-validity simulation

To make explicit what the neural contribution fraction, already framed in Methods as a divergence-based index rather than a validated measure of authorship or causal contribution, does and does not respond to, A dedicated simulation constructs six hand-specified pairs of 36-way distributions over the grid alphabet and computes the fused posterior and NCF for each using the same fuse and NCF formula (S1.4) as the primary analysis: sources that agree, sources that confidently conflict, a near-coin-flip neural signal tipped by a confident prior, an extremely confident neural signal against a comparatively diffuse prior, an uninformative (uniform) neural signal, and a neural signal that is overconfident relative to its true discriminability. This simulation is deterministic and independently reproducible; the values are reported in eTable 12.

#### S1.10 Temperature-scaling sensitivity

As described in Methods (Fusion and Posterior Calibration), both co-primary measures were also recomputed under an unfitted grid that raises p_neural and p_lm to the power 1/T and renormalizes each distribution before fusion. T < 1 sharpens each distribution toward its own arg max; T > 1 flattens it toward uniform. The join reconstructing raw probability vectors per selection follows the same (prior, context-prefix) lookup used to build the primary attribution frame. Values are reported in eTable 13.

This grid is a diagnostic sensitivity check, not a calibration procedure: T is chosen arbitrarily from a small exploratory set, applied identically to both sources regardless of whether either is actually over- or under-confident, and never fit against the true target symbol. It answers whether the co-primary estimates are sensitive to an arbitrary, joint rescaling applied the same way to both sources at once, holistically, not whether either source's own confidence is miscalibrated. Supplementary Note S1.10b below reports a separate procedure that instead fits an independent temperature to each source's own posterior using held-out data and the true target symbol, correcting that source's own calibration specifically; the two analyses answer different questions, and neither substitutes for the other.

#### S1.10b Independently-calibrated fusion (held-out, per-source temperature)

The per-source, held-out temperature-fitting procedure is described in Methods (Fusion and Posterior Calibration). Selections were grouped by participant into 5 folds; for each fold, the neural and language-model temperatures were fit only on the other 4 folds, then applied to raise and renormalize that source's posterior for the held-out fold's own selections, so no selection's calibrated posteriors were ever informed by that selection's own participant's data.

The reported 95% confidence intervals on the co-primary calibrated estimates come from resampling participants and recomputing the fused outcome on their already-calibrated posteriors; the fold temperatures themselves are fit once on the full data (grouped by participant, as above) and are not independently refit inside each bootstrap replicate. The intervals therefore reflect uncertainty in the fused outcome given this calibration, not uncertainty in the calibration fit itself.

Because T is fit to held-out data separately for each source rather than swept over an arbitrary shared grid, this procedure corrects each source's own over- or under-confidence rather than testing sensitivity to a joint rescaling: it is a calibration, not a sensitivity check, and it is not interchangeable with S1.10. Values are reported in eTable 21.

#### S1.11 Alternative attribution measures

The neural contribution fraction (S1.4) formalizes "how much of the fused posterior is attributable to the prior" as a Kullback-Leibler divergence between the fused posterior and each source's own distribution. Two non-KL alternatives were computed at the primary prior and beta = 1 to test whether the qualitative finding replicates under different formalisms rather than reflecting this specific measure.

**Log-odds contribution.** With n the neural posterior, p the prior, and k the intended symbol, each source's own conviction is its log-probability on k relative to the uniform baseline:

neural-shift = log n(k) - log(1/36); prior-shift = log p(k) - log(1/36)

The prior's share of the combined shift is prior-shift / (neural-shift + prior-shift). Unlike the neural contribution fraction, this measure looks only at the target symbol's odds under each source independently, never at the fused posterior itself, and is undefined (returned as NaN) only when the two shifts sum to exactly zero.

**Two-player Shapley value.** The fused posterior is treated as the outcome of a two-player coalition game with players "neural" and "prior." The coalition value function is the log-probability the coalition assigns the intended symbol: v(∅) = log(1/36), v({neural}) = log n(k), v({prior}) = log p(k), v({neural, prior}) = log f(k). With exactly two players the exact Shapley value reduces to averaging each player's marginal contribution to the coalition value across both join orders:

neural-Shapley = 0.5 × [v({neural}) - v(∅)] + 0.5 × [v({neural, prior}) - v({prior})] prior-Shapley = 0.5 × [v({prior}) - v(∅)] + 0.5 × [v({neural, prior}) - v({neural})]

By the efficiency property of the Shapley value, neural-Shapley + prior-Shapley always equals the total coalition gain v({neural, prior}) - v(∅). The prior's normalized share is prior-Shapley / (neural-Shapley + prior-Shapley). Values are reported in eTable 14.

#### S1.12 Context-permutation negative control

Each selection's neural evidence was fused, at the primary prior (Qwen2.5-32B) and beta = 1, against that prior's own already-scored p_lm for a context-prefix other than its true one, as a permutation negative control testing whether attribution to the prior reflects genuine linguistic relevance to the intended word rather than generic posterior sharpening from any sufficiently confident prior. The permutation reassigns which context-prefix each selection is joined against, with a fixed seed (20260802), requiring no new language-model inference. Because 662 of the 3,373 selections share an identical context-prefix (the empty string, at the first character of a word), a permutation that only avoids mapping each row onto its own index can still map it onto a different row carrying identical context text; the implementation instead groups rows by context-prefix value and rotates the group order by an offset at least as large as the largest group, guaranteeing that every selection receives a context-prefix that differs from its own true context while drawing from exactly the same multiset of observed contexts (a permutation, not resampling with replacement). Values are reported in eTable 15.

#### S1.13 Neural and prior source calibration diagnostics

The Bayesian fusion rule (S1.4) combines the neural posterior and the language-model prior multiplicatively. This is principled only if the neural posterior does not already carry an implicit prior over symbol frequency that would then be counted a second time once the explicit language-model prior is multiplied in. The flash-level logistic classifier underlying the neural posterior (S1.2) is fitted on EEG epoch features alone and never receives symbol identity as an input, so it has no way to learn a prior over which symbol is the target from natural letter or digit frequency; this is a structural property of the classifier's inputs, not a consequence of the balanced class weighting used to fit it. The 36-way neural posterior, built by accumulating this classifier's flash log-odds onto whichever symbols each flash illuminated and normalizing (the evidence-accumulation formula in S1.2), inherits that symbol-agnosticism directly: it cannot favor any symbol a priori by natural frequency, regardless of how the underlying classifier was trained. Multiplying it by an explicit, non-uniform language-model prior therefore does not double-count a natural-frequency prior, provided neither term is renormalized in a way that reintroduces natural symbol frequency into the neural term a second time.

Balanced class weighting instead bears on a distinct property: whether the classifier's own confidence is well calibrated. Because a single flash illuminates only a subset of the grid rather than the intended symbol alone, in every stimulation condition in this archive a given flash is far more often non-target than target for the selection in progress (checkerboard flashes illuminate 3 to 5 symbols, row-column flashes a full row or column of 6, S1.1), a stimulation-geometry artifact unrelated to symbol identity; without balanced weights, the classifier's output could be shaped by that flash-level class imbalance rather than by genuine discriminability. Even a classifier with no access to symbol identity could still distort the fused posterior if its confidence were systematically miscalibrated, because log-odds accumulated across many flashes would over- or under-weight the neural term relative to the prior irrespective of which symbol they favored. This is a consistency argument for symbol-neutrality, not a proof that the neural posterior's confidence is itself well calibrated, which is an empirical question answered directly below.

Two calibration measures are computed over the neural posterior alone at the primary prior's selection set (n = 3,373), independent of the language-model prior and the fusion rule, : a target-probability variant and a top-label variant (defined below). Both use the same binning procedure: 10 equal-width bins on whatever is passed in as "confidence," each bin's selection-count-weighted absolute gap between mean confidence and mean empirical accuracy, so the two measures differ only in what is bound to "confidence" and checked against correctness, not in how binning itself is done.

The target-probability variant is computed as follows. Its Brier score is the mean squared error between the full 36-way neural posterior and a one-hot vector at the intended symbol; it is 0 for a posterior that places all mass on the intended symbol and rises toward a maximum of 2 as probability mass is placed on the wrong symbols with increasing confidence. Its ECE uses the probability the neural posterior places on the intended symbol (not the posterior's own top choice) as "confidence," checked against whether the neural posterior's own arg max equals the intended symbol. Binning on P(true label) rather than on the classifier's own top probability is a nonstandard, fusion-motivated choice: it is exactly the quantity multiplied into the fused posterior, so it is the calibration diagnostic the Bayesian fusion rule directly depends on, but it answers a different question from the standard top-label ECE computed next. Across the 3,373 selections, this measure's Brier score was 0.438 and its ECE was 3.9% across 10 bins.

The standard top-label variant is computed as follows. Confidence is max(p_neural), the neural posterior's own top probability over all 36 symbols, checked against the same correctness label used above. This is the calibration question usually meant by "is the classifier calibrated" (when the classifier is N% confident in its own top choice, is it right N% of the time), and it is the measure a reviewer expecting standard multiclass ECE would look for. Its Brier score is the mean squared error between max(p_neural) and the binary correctness indicator, this is on a 0-to-1 scale, not the 0-to-2 scale of the 36-way target-probability Brier score above, so the two Brier values are not directly comparable in magnitude. Across the same 3,373 selections, mean top-choice confidence was 88.5% against 71.8% empirical accuracy: Brier score 0.171 and ECE 16.7% across 10 bins.

The two measures disagree substantially (3.9% versus 16.7% ECE) because they bin different events that coincide only on already-correct selections. When the neural posterior's arg max is wrong, its probability on the true label is typically low (appropriately signaling low confidence in that bin), while its probability on its own (wrong) top choice can still be high (signaling confidence the top-label measure then penalizes against 0% accuracy in that bin). Neither measure alone establishes whether the neural posterior's confidence is trustworthy in an absolute sense, only that the answer depends on which quantity is being asked about; neither addresses the language-model prior's own calibration, nor whether either source's pre-scaling confidence is corrected by the same held-out temperature calibration used for this study's principal co-primary estimates (Supplementary Note S1.10b). Both questions are addressed directly below.

This diagnostic generalizes the standard top-label definition above to any single source's probability distributions, reporting three measures side by side rather than one: the same full-distribution Brier score as the target-probability variant above (0-to-2 scale), the same standard top-label ECE as the top-label variant above (confidence = max probability, correctness = arg max matches the intended symbol, 10 equal-width bins), and the mean negative log-likelihood of the intended symbol under the source's own distribution , the quantity the held-out temperature calibration in Supplementary Note S1.10b directly minimizes. This combination (full-distribution Brier paired with top-label ECE) is the more conventional pairing in the calibration literature; the neural-only figures above were split into two separate "variants" specifically because each bound Brier and ECE to the same confidence definition, for a direct within-measure comparison, not because either used a nonstandard Brier formula. Applied to the neural posterior's pre-scaling (T = 1) distributions, it reproduces the Brier score (0.438) and ECE (16.7%) already given above: the same two numbers, recombined, not new values.

The same three measures are computed for all four combinations of source (neural posterior, language-model prior) and basis (pre-scaling raw posteriors at T = 1; post-scaling posteriors from the held-out, 5-fold per-source temperature calibration of Supplementary Note S1.10b and eTable 21), and reported in full in eTable 24. Pre-scaling, both sources were markedly miscalibrated by the standard top-label measure (Methods), consistent with both requiring substantial held-out temperature flattening to fit (T approximately 3.1 for the neural posterior, T approximately 2.4 for the prior; eTable 21). After applying each source's own held-out-fit temperature, ECE fell for both (Methods), alongside parallel drops in NLL (neural: 2.091 to 1.189; prior: 3.836 to 3.253) and Brier score (neural: 0.438 to 0.392; prior: 1.111 to 0.951). Every measure, for both sources, improved after scaling; none worsened.

This improvement is the direct, expected consequence of fitting each source's temperature against held-out mean negative log-likelihood (Supplementary Note S1.10b) and then evaluating that same fitted temperature's effect on Brier and ECE, not an independent confirmation from an unrelated diagnostic. It also does not, by itself, establish that either source is calibrated in an absolute sense: no threshold for "well calibrated" is adopted here, the post-scaling residual (4.1 to 4.8% ECE) is not zero, and Brier scores well above their respective floors (0 for a source that places all mass correctly) indicate that a meaningful amount of miscalibration remains for both sources even after correction. What the four-cell comparison does establish is that the held-out per-source temperature calibration underlying this study's principal co-primary estimates (Results) measurably improves, by every standard diagnostic checked here, the trustworthiness of both sources' confidence relative to the raw pre-scaling posteriors on which the earlier, uncalibrated sensitivity comparison relies (eTable 13); it does not certify either source, or the fused result, as calibrated outright.

Neither this comparison nor the neural-only measures above establishes that the neural and language-model terms are correctly scaled *relative to each other* for multiplicative fusion: that each source's own confidence is well-behaved does not imply the two are weighted correctly against one another once fused. Per-source temperature scaling corrects each source's own confidence independently and says nothing about their relative weight; that separate question is only partially addressed by the fusion-exponent sensitivity analysis (eTable 6), which shows the primary analysis used beta = 1 because it was the prespecified, unweighted exponent, not one calibrated against held-out data (Discussion, main text).

#### S1.14 Closed-loop emitted-context sensitivity

The primary analysis conditions the language-model prior on the archive's intended preceding characters rather than on the fused rule's own emissions (Methods, Language-Model Priors; S1.3), which isolates attribution from error propagation, a separate phenomenon in which an earlier wrong emission changes the context available to a later selection in the same word. This analysis repeats the pipeline with emitted context instead: for each of the 662 phrases (one per source recording, the same unit at which context-prefix resets to the empty string in the primary analysis, S1.3), selections are walked in order and the primary prior (Qwen2.5-32B) is scored, using the identical prefix-locked marginalization (S1.3), against the characters the fused decision rule itself emitted at earlier positions in the same phrase rather than the archive's ground truth; the score is fused with the same, already-computed neural posterior (S1.2) at beta = 1, and the resulting emission is appended to that phrase's running context before scoring the next selection. Because every selection can therefore carry a unique, session-specific context that cannot be scored once per shared string and reused in parallel the way the rest of the ladder is, this analysis required a dedicated sequential GPU job, run once for the primary prior only. Values are reported in eTable 16.

#### S1.15 Excluding dynamic stopping with bigram model

The archive's condition field (S1.1) distinguishes eight recorded stimulation and acquisition conditions (eTable 8). One, labeled DynBigram ("dynamic stopping with bigram model" in eTable 8), ran a bigram language-model prior online during the original session to decide when to stop flashing, rather than following a fixed flash sequence as every other condition did. Its 348 selections are therefore not independent of a language prior in the way this study's design assumes: the archived selection could already reflect a language-informed decision made before this study's own, offline language-model prior was ever applied to it, so its attribution risks being circular. "Dynamic stopping" without the bigram model (Dyn, 359 selections) also stops a trial early, but does so using only the neural classifier's own confidence; no other condition label in the archive documents an online adaptive or language-informed stopping rule (eTable 8), so DynBigram is the only condition this concern applies to.

This sensitivity analysis removes the 348 DynBigram selections from the primary-prior frame and recomputes both co-primary estimates, as participant-clustered bootstrap means (S1.6). It is run twice, once on the raw, uncalibrated fusion basis and once on the calibrated frame (S1.10b), so the check is available on the same basis the co-primary estimates are reported on. The calibrated run filters the already-calibrated frame rather than refitting the held-out temperatures on the 3,025-selection remainder, so each excluded selection still contributed to the fold temperature its retained neighbours were scored with. Those temperatures are two shared, per-source scalars fit over thousands of selections, so that residual contribution is diluted, though the calibrated row remains a filtered-frame estimate rather than a full recalibration. The same applies to the calibrated form of the pooled-calibration-session exclusion (eTable 8). Values are reported in eTable 17.

#### S1.16 Architecture-family permutation test and scale-equivalence test

The Spearman correlations in eTable 9 test whether attribution trends with log parameter count, but a null correlation across 21 points does not by itself distinguish a true absence of a scale effect from a study too small to detect one. Two formal tests address this directly. First, an architecture-family permutation test tests whether architecture family explains any of the variance in each attribution measure beyond chance: it operates on the same per-prior, participant-weighted mean table used elsewhere in this analysis (Methods), shuffles the 21 priors' family labels 10,000 times with a fixed seed, and on each shuffle recomputes a one-way-ANOVA-style between-family/within-family variance ratio (an F-like statistic that is always non-negative, so its one-sided upper-tail permutation count is already the two-sided P value). The observed statistic is compared against this permutation null.

Second, a two-one-sided-tests (TOST) equivalence test for the Spearman rho between log parameter count and each attribution measure, against a predefined margin of rho = ±0.3, a conventional small-effect threshold fixed before this analysis was run and not derived from the data. The cluster bootstrap (Methods, Statistical Analysis) resamples participants and does not apply to a per-prior correlation, so this test instead resamples the 21 per-prior points with replacement (same seed) and recomputes rho on each of 2,000 bootstrap replicates. The resulting 90% CI is the interval appropriate for a two-one-sided-tests equivalence conclusion at the conventional two-sided 0.05 level: equivalence is concluded only when that interval falls entirely within the predefined margin.

Both tests were run for all four attribution measures. Values are reported in eTable 18.

#### S1.17 Prior ladder growth

The prior ladder grew in five stages, tracked through the git commit history of the analysis repository. An initial six-tier ladder (uniform, character 5-gram, GPT-2, GPT-2-large, Qwen2.5-1.5B, Qwen2.5-3B) was scored first, using the co-primary outcome definitions given in S1.4 throughout; those definitions did not change at any later stage.

That six-tier ladder showed a trend between phantom agreement and parameter count, and the ladder was extended the same day (adding Qwen2.5-14B, Qwen2.5-32B, and Qwen3.5-27B) specifically to test whether the trend would hold. Qwen2.5-32B, the largest model in the resulting nine-entry ladder, was designated the primary prior at that point. The trend did not hold: it weakened rather than strengthened as more of the parameter range was sampled, a truncation-artefact pattern reported in Results (Effect of Parameter Count) and Discussion and confirmed by the ladder's later growth (eTable 9).

Three further extensions brought the ladder to its final 21 neural priors across eleven architecture families, alongside the three classical 5-grams: a cross-family round adding Llama, Gemma, Mistral, Mixtral, and DeepSeek priors and two additional n-gram variants; a second round filling a within-family statistical gap for the Gemma family; and a systematic architecture search using a blinded loadability preflight (eTable 10) that added Pythia, Mamba, and RecurrentGemma. Outcome definitions were unchanged throughout these three rounds.

### S2 Supplementary Tables

**eTable 1. Selection eligibility and exclusions.** Counts describe the path from archived recordings to the analysis frame. Trial-level exclusions removed selections whose feedback phase was not preceded by a stimulation window and selections retaining fewer than two artefact-free flash epochs; 2 recordings retained no selection on those grounds.

| Stage | Value |
| --- | --- |
| Test-phase recordings in the four ALS source studies | 667 |
| Recordings excluded, target code outside the 36-symbol grid | 1 |
| Recordings excluded, per-symbol flash channel absent | 2 |
| Recordings retaining no eligible selection after trial-level exclusions | 2 |
| Recordings contributing at least one analysed selection | 662 |
| Eligible selections in the analysis frame | 3,373 |
| Sessions | 115 |
| Participants | 47 |
| Sessions using pooled participant calibration | 2 |
| Flash epochs excluded for artefact | 3.4% |
| Selections with undefined neural contribution fraction | 0 |

**eTable 2. Attribution by character position within the intended word, raw fusion (sensitivity comparison to eTable 28).** Position 1 is the first character of a word, where the prior has no within-word context. Estimates are participant-weighted with 95% confidence intervals from 2,000 participant-cluster bootstrap replicates. eTable 28 reports the calibrated-basis counterpart, the primary basis for the Results' Secondary Outcomes section.

| Position | Selections | Phantom agreement, % | 95% CI | Neural contribution fraction | 95% CI |
| --- | --- | --- | --- | --- | --- |
| 1 | 662 | 2.4 | 1.6, 3.2 | 0.939 | 0.923, 0.959 |
| 2 | 662 | 3.5 | 2.3, 4.6 | 0.936 | 0.920, 0.952 |
| 3 | 661 | 3.5 | 2.4, 4.5 | 0.920 | 0.900, 0.942 |
| 4 | 542 | 3.7 | 2.3, 5.0 | 0.916 | 0.895, 0.939 |
| 5 | 429 | 7.0 | 5.1, 8.7 | 0.879 | 0.850, 0.909 |
| 6 | 417 | 6.5 | 4.0, 8.8 | 0.894 | 0.869, 0.919 |

Two features of this design could confound position with something other than position itself: the same word recurring across the corpus, and clustering within a recording session; and the fact that words of different lengths reach different maximum positions, so "later position" is not fully separable from "closer to the end of a shorter word." Both are addressed directly in eTable 20.

**eTable 3. Attribution and reconstruction fidelity by calibration decoder quality, raw fusion (sensitivity comparison to eTable 28).** Tertiles are formed on grouped cross-validated calibration area under the curve. Fidelity is the proportion of selections whose offline posterior argmax matched the selection recorded online in the archive. eTable 28 reports the calibrated-basis attribution counterpart (fidelity itself has no calibrated analog, since it compares the offline posterior's argmax to the archive's online selection regardless of fusion basis).

| Tertile | Selections | Phantom agreement, % | 95% CI | Neural contribution fraction | Reconstruction fidelity, % |
| --- | --- | --- | --- | --- | --- |
| Low | 1,165 | 5.8 | 4.4, 7.0 | 0.860 | 39.2 |
| Middle | 1,087 | 3.3 | 2.1, 4.5 | 0.951 | 80.5 |
| High | 1,121 | 2.5 | 1.4, 3.5 | 0.970 | 90.6 |

**eTable 4. Attribution by target type, raw fusion (sensitivity comparison to eTable 28).** Digit-string targets were the prespecified low-predictability stratum. eTable 28 reports the calibrated-basis counterpart.

| Target type | Selections | Phantom agreement, % | 95% CI | Neural contribution fraction |
| --- | --- | --- | --- | --- |
| English word | 3,205 | 3.7 | 3.0, 4.4 | 0.924 |
| Digit string | 168 | 6.1 | 1.4, 10.2 | 0.925 |

**eTable 5. Secondary-family hypothesis tests with multiplicity correction, raw fusion (sensitivity comparison to eTable 30).** The two continuous rows are the linear mixed model with a participant random intercept, which is the sensitivity comparison for those relationships and the source of this outcome's adjusted P values; the bounded fractional-logit model summarized as predicted-value contrasts is primary for them and is fit on the calibrated basis only, so it appears in eTable 30 and has no counterpart here (Supplementary Note S1.6). Binary outcomes use logistic generalized estimating equations clustered on participant. Adjusted P values use the Benjamini-Hochberg procedure across the five-member family. eTable 30 reports the calibrated-basis counterpart, the primary basis for the Results' Secondary Outcomes section.

| Test | Estimate | 95% CI | Raw P | Adjusted P |
| --- | --- | --- | --- | --- |
| Phantom agreement per character of context, odds ratio | 1.18 | 1.05, 1.31 | 0.004 | 0.004 |
| Phantom agreement per unit calibration AUC, odds ratio | 0.042 | 0.008, 0.217 | <0.001 | <0.001 |
| Phantom agreement, digit versus word target, odds ratio | 1.53 | 0.64, 3.64 | 0.338 | 0.338 |
| Neural contribution fraction per character of context, slope | -0.0067 | -0.0100, -0.0033 | <0.001 | <0.001 |
| Neural contribution fraction per unit calibration AUC, slope | +0.545 | +0.451, +0.639 | <0.001 | <0.001 |

**eTable 6. Sensitivity to the fusion exponent.** The exponent weights the neural term relative to the prior. A value of 1 is the standard Bayesian product and the primary analysis.

| Fusion exponent | Phantom agreement, % | Prior capture, % | Neural contribution fraction |
| --- | --- | --- | --- |
| 0.25 | 5.7 | 13.7 | 0.434 |
| 0.5 | 5.5 | 5.4 | 0.797 |
| 1 | 3.8 | 1.8 | 0.925 |
| 2 | 2.2 | 0.7 | 0.959 |
| 4 | 1.4 | 0.3 | 0.963 |

**eTable 7. Leave-one-study-out sensitivity, raw fusion (calibrated counterpart in eTable 32).** Each row removes one source study and recomputes both co-primary outcomes on the remainder.

| Study removed | Phantom agreement, % | Neural contribution fraction |
| --- | --- | --- |
| Study B | 5.3 | 0.892 |
| Study F | 3.5 | 0.924 |
| Study L | 3.5 | 0.928 |
| Study N | 3.1 | 0.946 |

**eTable 8. Attribution by stimulation condition, raw fusion (calibrated basis in eFigure 2).** Conditions are those recorded in the archive. The bigram condition ran a language prior online during the original session.

| Condition | Selections | Phantom agreement, % | Neural contribution fraction |
| --- | --- | --- | --- |
| Static | 360 | 0.8 | 0.970 |
| Checkerboard | 1,176 | 3.2 | 0.951 |
| Checkerboard column | 330 | 3.3 | 0.928 |
| Row-column | 330 | 4.2 | 0.902 |
| Dynamic stopping with bigram model | 348 | 6.4 | 0.905 |
| Wet electrodes | 234 | 6.5 | 0.852 |
| Dynamic stopping | 359 | 6.7 | 0.902 |
| Dry electrodes | 236 | 7.6 | 0.785 |

Excluding the 2 sessions that used pooled participant calibration left phantom agreement at 3.8% and the neural contribution fraction at 0.925 across the remaining 3,299 selections on this raw basis, and at 4.5% and 0.914 on the calibrated basis, each within 0.1 percentage points or 0.001 of the corresponding full-cohort estimate. The calibrated form filters the already-calibrated frame rather than refitting the held-out temperatures on the remainder (Supplementary Note S1.15).

**eTable 9. No monotonic association with parameter count or architecture family was detected, raw fusion (sensitivity comparison to eTable 23).** Spearman rank correlations between each attribution measure and the base-10 logarithm of parameter count, across the 21 neural language models spanning eleven architecture families (GPT-2, Qwen, Llama, Gemma, Mistral, DeepSeek, OLMo, GPT-OSS, Pythia, Mamba, RecurrentGemma), computed on raw, uncalibrated fusion. The final two columns repeat each correlation on the ladder truncated below 10 billion parameters (11 of the 21 neural language models), the closest analog to the original single-family truncation. eTable 23 reports the calibrated-basis counterpart of this table, the primary basis for the Results' Effect of Parameter Count section.

| Measure | Range across the ladder | rho, full ladder | P, full ladder | rho, truncated <10B | P, truncated <10B |
| --- | --- | --- | --- | --- | --- |
| Phantom agreement | 2.5 to 4.9% | +0.11 | .63 | +0.62 | .044 |
| Neural contribution fraction | 0.907 to 0.944 | +0.08 | .72 | -0.54 | .088 |
| Prior capture | 1.2 to 3.3% | -0.10 | .65 | +0.45 | .16 |
| Accuracy gained by adding the prior | -0.3 to 2.5 points | +0.22 | .34 | +0.13 | .71 |

In the earlier, sparser 13-prior ladder, the truncated-range correlations for prior capture and accuracy gained reached conventional significance (*P* = .007 and .01) while the full-ladder correlations were null. With 11 rather than 9 priors now filling the same truncated range and the corrected scorer in place, neither of those two correlations remains significant (prior capture, *P* = .16; accuracy gained, *P* = .71). Instead, phantom agreement's truncated-range correlation now reaches marginal significance (rho = +0.62; *P* = .044), where it did not before the correction; this is a single, small-sample (n = 11) exploratory correlation, uncorrected for the four measures tested here, and its full-ladder counterpart remains null (rho = +0.11; *P* = .63). Which measure shows a truncated-range signal has changed across successive ladder extensions and this scorer correction, itself further evidence that these narrow-range correlations are unstable rather than reflecting a real parameter-count effect. All four full-ladder correlations remain null, so the apparent parameter-count effect continues to be a property of the sampled range rather than of parameter count itself.

**eTable 10. Architecture search and prior-model inclusion (cutoff 2026-08-01).** A search for additional causal-language-model architectures identified 6 candidates spanning families not yet represented in the ladder. Each candidate's exact checkpoint and tokenizer revision was frozen before scoring; inclusion required passing a compute-cluster preflight (a 31-context smoke test requiring finite, normalized 36-way priors and a byte-identical deterministic repeat) followed by the same 951-context scoring used for every other prior. No candidate was included or excluded on the basis of its observed attribution result.

| Candidate | Family | Parameters | Architecture (arXiv) | Outcome | Reason |
| --- | --- | --- | --- | --- | --- |
| RecurrentGemma-2B | RecurrentGemma | 2.7B | Griffin: gated linear recurrence plus local sliding-window attention (2402.19427) | Included | Passed preflight and full 951-context scoring; results in Table 2, Figure 3, eTable 9 |
| RWKV-7 "Goose" 2.9B | RWKV | 2.9B | RWKV-7 generalized delta-rule linear attention (2503.14456) | Excluded | Checkpoint weights for 6 attention parameters per layer did not match the installed transformers library's native parameter names and were randomly initialized rather than loaded; the deterministic-repeat gate correctly failed on both attempts |
| xLSTM-7B | xLSTM | 7B | Recurrent memory with modern sLSTM/mLSTM cells (2503.13427) | Excluded | Checkpoint config requests an unsupported kernel variant that the installed model-loading library's config validator does not accept; failed on both attempts |
| StripedHyena-Hessian-7B | StripedHyena | 7B | Hybrid attention plus gated Hyena convolution (2302.10866) | Excluded | Requires the FlashAttention package, unavailable as a prebuilt wheel for the installed CUDA/PyTorch/Python combination; declined an unverified from-source compile |
| RetNet 2.7B | RetNet | 2.7B | Retentive Network: parallel, recurrent, and chunkwise multi-scale retention (2307.08621) | Excluded | Checkpoint weights loaded correctly, but the deterministic-repeat gate failed on both attempts (Triton kernel backend flagged the installed Triton version as below the library's recommended minimum) |
| Gated DeltaNet 1.3B | Gated DeltaNet | 1.3B | Gated delta-rule linear-attention stack (2412.06464) | Excluded | Checkpoint weights loaded correctly, but the deterministic-repeat gate failed on both attempts (same Triton kernel backend as RetNet) |

**eTable 11. Neural support among phantom-agreement selections, raw fusion (sensitivity comparison to eTable 26).** For every selection where the fused posterior selected the intended character but the neural posterior alone would not have (n = 142), the neural probability assigned to the intended character and its rank within the 36-symbol neural posterior. A phantom-agreement selection with substantial neural probability on the intended character (the "0.49 vs 0.51" case) is a different phenomenon from one where the neural evidence assigned it almost no probability at all; both currently register identically as "phantom agreement," and this table reports the split. eTable 26 reports the calibrated-basis counterpart, the primary basis for Figure 4 and the Results paragraph beside it.

| Neural probability on intended character | Share of phantom-agreement cases |
| --- | --- |
| Below 0.10 | 34.5% |
| 0.10 to 0.40 | 55.6% |
| 0.40 or above | 9.9% |

| Neural rank of intended character | Share of the 137 untied cases |
| --- | --- |
| Second-ranked (neural's runner-up) | 66.4% |
| Third-ranked | 21.2% |
| Fourth-ranked or worse | 12.4% |

Median neural probability on the intended character: 0.171. Median margin between the neural posterior's own top choice and the intended character: 0.394. The rank breakdown omits 5 cases (3.5%) in which the intended character was tied with the neural posterior's own top-probability symbol (rank 0); a phantom-agreement label is still possible in this situation because the deterministic tie-break used to convert the neural posterior into a single decoded symbol did not resolve to the intended character even though its probability was not exceeded by any other symbol.

**eTable 12. What the neural contribution fraction does and does not distinguish.** Six synthetic scenarios (Supplementary Note S1.9) isolate what the Kullback-Leibler-based measure responds to. It is a divergence-based index of how far the fused posterior moved from each source, not a validated measure of authorship, causal contribution, or evidential responsibility; the miscalibrated scenario shows it cannot detect an overconfident neural source from the distributions alone.

| Scenario | Neural contribution fraction | What it shows |
| --- | --- | --- |
| Agreement | 0.500 | Both sources point to the same symbol; is not exclusively attributable to either. |
| Conflict | 0.500 | Confident sources disagree; because they are equally confident, the displacement splits evenly regardless of which symbol is ultimately favored. |
| Near tie | 0.129 | A near-50/50 neural signal is easily tipped by a confident prior. |
| Sharply peaked neural | 0.997 | An overwhelming neural signal dominates a comparatively diffuse prior. |
| Diffuse neural | <0.001 | An uninformative (uniform) neural posterior yields full prior determination by construction. |
| Miscalibrated neural | 0.847 | An overconfident neural source is indistinguishable, by this measure alone, from a genuinely discriminative one. |

**eTable 13. Sensitivity of both co-primary measures to temperature scaling of the neural and language-model distributions (Supplementary Note S1.10).** Both sources were raised to the power 1/T and renormalized before fusion, at the primary prior and beta = 1.

| Temperature | Neural contribution fraction | Phantom agreement |
| --- | --- | --- |
| T = 0.5 (sharpened) | 0.914 | 4.2% |
| T = 1.0 (unscaled) | 0.920 | 4.2% |
| T = 2.0 (flattened) | 0.934 | 4.2% |

Phantom agreement is unchanged across the grid (4.2% at every temperature tested) because it, like every other categorical attribution measure, depends only on each source's own arg max and on the arg max of the fused posterior; raising a distribution to a positive power and renormalizing preserves that ordering, so which symbol each source and the fusion favor does not change. The neural contribution fraction, a continuous Kullback-Leibler-based measure, is the one that moves with temperature, rising modestly from 0.914 to 0.934 as the grid runs from sharpened to flattened distributions.

Unlike the participant-weighted estimates reported elsewhere in this study (S1.6), the values in this table are unweighted means across the 3,373 selections; consequently the T = 1.0 row does not exactly reproduce the raw, uncalibrated fusion's participant-weighted estimates, reported in the main text as a sensitivity comparison rather than as the co-primary result (neural contribution fraction 0.925; phantom agreement 3.8%; Results, Sensitivity Analyses). Recomputed with participant weighting, the T = 1.0 row reproduces those raw-fusion estimates exactly, confirming that the small gap reflects the weighting convention rather than the temperature-scaling procedure itself. The main text's co-primary result is no longer the raw, T = 1.0 fusion at all: it is the independently-calibrated fusion reported in S1.10b and eTable 21, which uses a different, per-source-fit T rather than the shared grid tested here.

**eTable 14. The prior's share of attribution under three formalisms (Supplementary Note S1.11).** All three measures were computed on the same 3,373 selections at the primary prior (Qwen2.5-32B) and beta = 1. The mean and median follow eTable 13's convention of unweighted per-selection averages; the range reports the minimum and maximum value observed across those 3,373 selections. The neural contribution fraction is reported as 1 minus itself so that all three rows read as "the prior's share."

| Attribution measure | Basis | Prior's share, mean | Prior's share, median | Prior's share, range |
| --- | --- | --- | --- | --- |
| 1 - Neural contribution fraction | Kullback-Leibler divergence of the fused posterior from each source | 0.080 | 0.001 | 0.000 to 1.000 (bounded by construction) |
| Log-odds contribution | Each source's own log-odds on the target symbol, relative to uniform, compared independently of the fused posterior | 0.941 | 0.075 | -240.5 to 932.4 |
| Two-player Shapley value (normalized) | Exact Shapley value; coalition value is the log-probability of the target symbol under each coalition | 0.235 | 0.012 | -125.3 to 675.4 |

Different attribution formalisms agreed qualitatively that the prior often influenced the fused output, but did not yield a stable or uniquely interpretable numerical share. The log-odds contribution's mean no longer agrees with the other two measures on whether the prior's typical share is even a minority: the Kullback-Leibler-based measure's mean (0.080) and the exact Shapley value's mean (0.235) are both minority shares, while the log-odds contribution's mean (0.941) is not. This is a property of the measure's denominator, not evidence that the prior's typical contribution changed direction. In a minority of selections the neural and prior log-odds shifts nearly cancel (their sum approaches zero without reaching the 1e-12 threshold that returns an explicit NaN), which inflates the ratio to extreme values (log-odds contribution ranges from -240.5 to 932.4 across the 3,373 selections) that dominate an arithmetic mean; the measure's median (0.075), which is not sensitive to those outliers, is a minority share consistent with the other two measures' medians. The Shapley share is subject to the same denominator mechanism at smaller magnitude (mean 0.235 vs median 0.012, range -125.3 to 675.4), so its numeric distance from the Kullback-Leibler-based measure's mean should not be read as immunity from the instability described above. The neural contribution fraction shows a different pattern: it is bounded to [0, 1] by construction, so no denominator instability is possible, yet its median (0.001) is still far below its own mean (0.080), because the measure concentrates near "fully neural" (NCF near 1, prior's share near 0) for the large majority of selections with confident, unambiguous neural evidence, while a smaller subset of harder selections with more balanced posteriors pulls the mean up. No single statistic here should be read as the definitive prior share; the co-primary neural contribution fraction remains the prespecified primary measure, and these alternatives are reported as a robustness check on its qualitative conclusion, that the prior often shapes which symbol the fused posterior favors, rather than as a source of a replacement point estimate.

**eTable 15. Stratified context-permutation negative control, raw fusion (sensitivity comparison to eTable 29) (Supplementary Note S1.12).** Each selection's neural evidence was fused against the primary prior's own p_lm for a context permuted within the same character-position, source-study, and context-length-bucket stratum (200 permutations, seed 20260802), rather than its true context. Of 3,373 selections, 663 (mostly a word's first character, which by construction has no other same-stratum context to draw from) had no valid permutation partner and are excluded from both the real-context comparator and the permutation null below; the remaining 2,710 selections are used throughout this table. Values are unweighted means, as in eTable 13 and eTable 14.

| Context | Neural contribution fraction | Phantom agreement |
| --- | --- | --- |
| Real (true, matched) context | 0.915 | 4.6% |
| Permutation null, mean across 200 permutations (95% CI) | 0.923 (0.919 to 0.926) | 2.9% (2.5 to 3.3%) |

The real context's phantom agreement (4.6%) exceeded every one of the 200 permutation draws (one-sided exceedance *P* < .005) and its neural contribution fraction (0.915) fell below every draw (*P* < .005), so both measures moved in the direction predicted if attribution to the prior reflects genuine linguistic relevance to the intended word: a real but wrong context supplies less useful evidence than the true context, so it less often determines a correct emission and the fused posterior sits closer to the neural evidence's own distribution. Every permuted context remains a real, confidently peaked language-model output rather than a uniform or degenerate one, so this result also weighs against an alternative explanation in which any sufficiently confident prior, regardless of relevance, would produce the same phantom-agreement rate.

**eTable 16. Closed-loop emitted-context sensitivity for the primary prior, raw fusion (sensitivity comparison to eTable 27) (Supplementary Note S1.14).** Each selection's neural evidence was fused against the primary prior scored on the fused decision rule's own emitted prefix within the same phrase, rather than the archive's intended prefix, and compared against the same selections fused against their true intended context, both under the same raw, uncalibrated (beta = 1) fusion rule, reported here as the disclosed sensitivity comparison; eTable 27 reports the calibrated-basis paired comparison, the primary basis for the Results paragraph beside it. The emitted (closed-loop) context row reports the participant-weighted bootstrap mean with 95% CI (2,000 participant-cluster bootstrap replicates, S1.6). The intended (archive) context row was not resampled for this comparison and remains the unweighted per-selection mean, as in eTable 13 through eTable 15; it is therefore not on the same statistical footing as the row below it, though the two point estimates are close.

| Context | Neural contribution fraction | Phantom agreement | 95% CI |
| --- | --- | --- | --- |
| Intended (archive) context, unweighted mean | 0.920 | 4.2% | not computed |
| Emitted (closed-loop) context, participant-weighted | 0.911 | 3.4% | NCF, 0.892 to 0.930; phantom agreement, 2.5% to 4.1% |

Of the 3,373 selections, 1,203 (35.7%) fell at a phrase position where at least one earlier selection in the same word had already been mis-emitted, so the emitted context differed from the intended context; the remaining 2,170 (64.3%) saw an emitted context identical to the intended one because every earlier character in that word had been emitted correctly. Unlike under the permuted-context negative control (eTable 15), the closed-loop condition did not move both metrics in the same direction relative to the intended-context estimate: phantom agreement fell (4.2% to 3.4%) while the prior's share of posterior displacement rose slightly rather than fell (8.0% to 8.9%). A wrong, error-compounding emitted prefix is not simply a weaker version of a permuted one: it remains internally consistent with the fused rule's own prior-influenced history within the same word, unlike a context drawn from an unrelated selection, which may explain why it does not reproduce the uniformly weaker-attribution pattern seen under permutation. The qualitative pattern under the two controls therefore diverges as well as differing in magnitude, so the primary, intended-context estimates should not be assumed to be a strict upper bound relative to emitted-context propagation in this offline reconstruction: the emitted-context row's 95% CI (2.5% to 4.1%) does not exclude the 3.8% (95% CI, 3.0 to 4.5) obtained for the intended context under the same raw fusion rule, computed with the same participant weighting and bootstrap procedure (Results, Sensitivity Analyses); it is this raw-fusion, intended-context estimate, not the calibrated co-primary estimate reported in Results and the Abstract, nor the 4.2% intended-context row shown above (an unweighted mean with no participant-clustered bootstrap CI computed for direct comparison), that is the appropriate comparator for this table.

**eTable 17. Excluding "dynamic stopping with bigram model," calibrated and raw fusion (Supplementary Note S1.15).** That condition, unlike every other one in eTable 8, ran a bigram language-model prior online during the original session, so its offline attribution could be circular: the archived selection had already been shaped by a language prior before this study's own prior was ever applied. This sensitivity removes its 348 selections and recomputes both co-primary measures, as participant-clustered bootstrap means (95% CI), on the remaining 3,025 of 3,373 selections. It is reported on both fusion bases: the calibrated basis the co-primary estimates in Results and the Abstract are computed on (Supplementary Note S1.10b), and the raw, uncalibrated (beta = 1) basis retained throughout as a labeled sensitivity comparison (Methods). The calibrated rows filter the already-calibrated frame rather than refitting the held-out temperatures on the remainder, a limitation described in Supplementary Note S1.15. Each full-cohort row is the corresponding co-primary estimate on its own basis.

| Cohort | Fusion basis | Selections | Neural contribution fraction | Phantom agreement, % |
| --- | --- | --- | --- | --- |
| Full cohort | Calibrated | 3,373 | 0.914 (95% CI, 0.896 to 0.934) | 4.4 (95% CI, 3.5 to 5.3) |
| Excluding dynamic stopping with bigram model | Calibrated | 3,025 | 0.917 (95% CI, 0.898 to 0.936) | 4.2 (95% CI, 3.4 to 5.1) |
| Full cohort | Raw | 3,373 | 0.925 (95% CI, 0.907 to 0.943) | 3.8 (95% CI, 3.0 to 4.5) |
| Excluding dynamic stopping with bigram model | Raw | 3,025 | 0.927 (95% CI, 0.909 to 0.945) | 3.6 (95% CI, 2.8 to 4.3) |

On both bases, both co-primary estimates were essentially unchanged, with 95% CIs that overlap almost entirely with the full-cohort estimates, so the online language prior present during these 348 selections' original acquisition is not driving either estimate. Among the archive's eight documented stimulation conditions (eTable 8), "dynamic stopping with bigram model" is the only one recorded as having used an online language-model prior; "dynamic stopping" without the bigram model (359 selections) also ends a trial early, but does so using only the neural classifier's own confidence, with no language model involved, and no other condition label in the archive implies an online adaptive or language-informed decision. Those 359 selections therefore carry no comparable circularity concern and were retained in this table's own remainder, as in every other analysis in this supplement.

**eTable 18. Formal tests of the parameter-count and architecture-family null result, raw fusion (sensitivity comparison to eTable 31) (Supplementary Note S1.16).** A permutation test for whether architecture family explains variance in each attribution measure beyond chance (family labels shuffled 10,000 times across the 21 neural language models, fixed seed), and a two-one-sided-tests (TOST) equivalence test for the Spearman correlation between log parameter count and each measure, against a predefined margin of rho = ±0.3 (21 priors resampled with replacement, 2,000 bootstrap replicates, same seed). eTable 31 reports the calibrated-basis counterpart of both tests, the primary basis for the Results' Effect of Parameter Count section.

| Measure | Permutation F-like statistic | Permutation *P* | Spearman rho | 90% CI | Equivalent at ±0.3? |
| --- | --- | --- | --- | --- | --- |
| Neural contribution fraction | 1.02 | .48 | +0.08 | -0.31 to 0.49 | No |
| Phantom agreement | 0.50 | .82 | +0.11 | -0.26 to 0.45 | No |
| Prior capture | 1.04 | .46 | -0.10 | -0.53 to 0.31 | No |
| Accuracy gained by adding the prior | 0.96 | .51 | +0.22 | -0.16 to 0.55 | No |

Neither test found an architecture-family or parameter-count effect the descriptive Spearman correlations in eTable 9 had not already suggested, and the permutation test found no measure's between-family variance exceeded chance. The equivalence test's bootstrap CIs, however, were wide enough that no measure formally confirmed equivalence at the predefined ±0.3 margin: with only 21 priors, resampling with replacement carries variability the point estimates alone do not convey. These results are therefore better read as a failure to reject a parameter-count or architecture-family effect than as positive evidence that none exists.

**eTable 19. Full prior-ladder inventory.** All 25 priors used in the primary analysis: a uniform null, three classical character 5-gram baselines, and 21 causal language models spanning eleven architecture families. Distinct from eTable 10, which documents architecture candidates that were screened but not all included; this table lists only priors actually scored and reported in Table 2.

| Prior | Family | Parameters | Type | Organization |
| --- | --- | --- | --- | --- |
| Uniform | Uniform | NA | NA | NA |
| Character 5-gram (add-k, backoff) | n-gram | NA | NA | NLTK 3.9.2 (Brown corpus) |
| 5-gram, Kneser-Ney | n-gram | NA | NA | NLTK 3.9.2 (Brown corpus) |
| 5-gram, Kneser-Ney (WikiText-103) | n-gram | NA | NA | HuggingFace datasets (WikiText-103) |
| GPT-2 | GPT-2 | 124M | Base | OpenAI |
| GPT-2 large | GPT-2 | 774M | Base | OpenAI |
| Qwen2.5-1.5B | Qwen | 1.54B | Base | Alibaba Cloud |
| Qwen2.5-3B | Qwen | 3.09B | Base | Alibaba Cloud |
| Qwen2.5-14B | Qwen | 14.7B | Base | Alibaba Cloud |
| Qwen2.5-32B | Qwen | 32.5B | Base | Alibaba Cloud |
| Qwen3.5-27B | Qwen | 27.0B | Instruct | Alibaba Cloud |
| Qwen3.6-35B-A3B | Qwen | 35.0B, total* | Instruct | Alibaba Cloud |
| Llama-3.1-8B | Llama | 8.0B | Instruct† | Meta |
| Gemma-2-2B | Gemma | 2.0B | Instruct | Google |
| Gemma-2-9B | Gemma | 9.0B | Base | Google |
| Gemma-2-27B | Gemma | 27.0B | Base | Google |
| Gemma-4-12B | Gemma | 12.0B | Instruct | Google |
| Mistral-7B | Mistral | 7.0B | Base | Mistral AI |
| Mixtral-8x7B | Mistral | 46.7B, total* | Base | Mistral AI |
| DeepSeek-V2-Lite | DeepSeek | 15.7B, total* | Base | DeepSeek |
| OLMo-2-7B | OLMo | 7.0B | Instruct | Allen Institute for Artificial Intelligence |
| GPT-OSS-20B | GPT-OSS | 20.9B, total* | Instruct | OpenAI |
| Pythia-12B | Pythia | 12.0B | Base | EleutherAI |
| Mamba-2.8B | Mamba | 2.8B | Base | Mamba authors' Hugging Face release |
| RecurrentGemma-2B | RecurrentGemma | 2.7B | Base | Google |

Fourteen of the 21 neural language models are base models and 7 are instruction-tuned. *Total parameters are shown for the mixture-of-experts or sparsely activated models (Qwen3.6-35B-A3B, Mixtral-8x7B, DeepSeek-V2-Lite, GPT-OSS-20B); active parameters per forward pass, as stated on each model's own card, are smaller: Qwen3.6-35B-A3B, 35B total and 3B active (Qwen's "A3B" suffix denotes 3B activated parameters); Mixtral-8x7B, 46.7B total and 12.9B active per token; DeepSeek-V2-Lite, 15.7B total and 2.4B active per token; GPT-OSS-20B, 20.9B total and 3.6B active per forward pass. †Substituted for a base Llama checkpoint because live Hugging Face Hub access to any Llama-family repository was not available to this account at scoring time despite the repositories showing as accessible in metadata queries (Methods). Neither Qwen3.6-35B-A3B nor Qwen3.5-27B has an arXiv technical report; each is cited in the main text to its own official model-card citation (Qwen Team, 2026), not to the unrelated Qwen2.5 Technical Report used for the four Qwen2.5 priors.

**eTable 20. The position-in-word effect after adjustment for word identity and session (both co-primary outcomes), and restricted to six-character words only (neural contribution fraction), raw fusion (Supplementary Note S1.6b). The first table below models the neural contribution fraction as a function of character position with a linear mixed model; the coefficient is the change in neural contribution fraction per additional character of context. The unadjusted row repeats the eTable 5 estimate for comparison. The word- and session-adjusted row adds word identity and recording session as crossed random effects alongside participant, fit with the lbfgs optimizer after the default optimizer failed to reach a reported convergence (Supplementary Note S1.6b); three further optimizers converged independently to the same coefficient to four decimal places. The six-character-words-only row reuses the unadjusted model's fitting call unchanged, restricted to the subset of selections whose intended word had exactly six characters, the only length reaching every position. The last table applies the same word- and session-adjustment to phantom agreement, which needs different machinery and is described in its own note directly above it. The calibrated-basis counterparts of every estimate here are reported in eTable 28.**

| Model | Selections | Position coefficient | 95% CI | P |
| --- | --- | --- | --- | --- |
| Unadjusted (eTable 5) | 3,373 | -0.0067 | -0.0100, -0.0033 | <0.001 |
| Word- and session-adjusted (crossed random effects) | 3,373 | -0.0068 | -0.0099, -0.0036 | <0.001 |
| Six-character words only | 2,502 | -0.0069 | -0.0107, -0.0031 | <0.001 |

The word- and session-adjusted coefficient is close to the unadjusted estimate, and the six-character-words-only coefficient is likewise close to it and in the same direction, slightly larger in magnitude. Neither adjustment attenuated the position effect; both findings are consistent with a genuine within-word position gradient rather than an artefact of recurring words, session clustering, or variable word length.

The crossed model's three variance components, on the neural-contribution-fraction scale, show that participant, session, and word identity all explained genuine, statistically distinguishable variance:

| Variance component | Variance | 95% CI | P |
| --- | --- | --- | --- |
| Participant | 0.155 | 0.039, 0.272 | 0.009 |
| Session | 0.130 | 0.066, 0.195 | <0.001 |
| Word identity | 0.066 | 0.028, 0.104 | 0.001 |

Phantom agreement, the binary co-primary outcome, carries its own position-in-word gradient and is adjusted for the same crossed word and session structure in the table below. The two rows come from different statistical machinery and are not interchangeable. The unadjusted row is the logistic generalized estimating equation reported for this outcome throughout the paper, which admits only one clustering variable and so cannot represent the crossed structure. The word- and session-adjusted row is a crossed-random-effects Bayesian mixed generalized linear model estimated by a variational approximation to the posterior (Supplementary Note S1.6b). Its interval is therefore a 95% credible interval from that approximate posterior rather than a sandwich-based confidence interval, and no P value is defined for it, which is why it is deliberately not a member of the Benjamini-Hochberg-adjusted secondary family (eTable 5) and carries no adjusted P value. The interval type is labeled per row for that reason.

| Model | Selections | Position odds ratio | 95% interval | Interval type | P |
| --- | --- | --- | --- | --- | --- |
| Unadjusted (eTable 5) | 3,373 | 1.18 | 1.05, 1.31 | Confidence | 0.004 |
| Word- and session-adjusted (crossed random effects) | 3,373 | 1.15 | 1.09, 1.21 | Credible | Not defined |

The adjusted odds ratio is close to the unadjusted one, in the same direction, and its interval excludes 1, so word identity and session do not account for phantom agreement's position gradient any more than they account for the neural contribution fraction's. The crossed model's three random-effect standard deviations, on the log-odds scale, are 0.32 for participant (95% credible interval, 0.26 to 0.39), 0.53 for word identity (0.49 to 0.57), and 0.25 for session (0.22 to 0.28); word identity is the largest of the three, so this is not an adjustment for a negligible source of clustering. No six-character-words-only refit is reported for phantom agreement, so word length is addressed for the neural contribution fraction only.

**eTable 21. Held-out, per-source temperature calibration and the resulting co-primary estimates (Supplementary Note S1.10b).** Selections were grouped by participant into 5 folds; each fold's temperatures were fit on the other 4 folds' selections only and applied to that fold's held-out selections. T > 1 indicates the source was overconfident on held-out data and was flattened toward uniform before fusion.

| Fold | Held-out selections, No. | T, neural posterior | T, language-model posterior |
| --- | --- | --- | --- |
| 1 | 673 | 3.07 | 2.40 |
| 2 | 672 | 2.92 | 2.38 |
| 3 | 670 | 3.12 | 2.40 |
| 4 | 682 | 3.29 | 2.41 |
| 5 | 676 | 3.20 | 2.39 |
| Mean across folds | 3,373 | 3.12 | 2.40 |

Both sources required substantial flattening in every fold, more for the neural posterior (range, 2.92 to 3.29) than the language-model posterior (range, 2.38 to 2.41), indicating the neural decoder was the more overconfident of the two sources on held-out data. Fusing the resulting calibrated posteriors at beta = 1 gave a neural contribution fraction of 0.914 (95% CI, 0.896 to 0.934) and phantom agreement of 4.4% (95% CI, 3.5 to 5.3), the co-primary estimates reported as the principal result in the main text (Results). These values are not directly comparable to eTable 13's T = 1.0 row: eTable 13 leaves both sources unscaled (T = 1.0 is the identity), while this table's T values are fit, per source, to correct each source's own confidence, and neither table's T is applied to the other.

**eTable 22. Attribution measures across the language-model prior ladder, raw fusion (sensitivity comparison to Table 2).** The same prior ladder as Table 2, computed on raw, uncalibrated fusion (beta = 1, T = 1 for both sources) rather than the held-out, per-source temperature calibration that is Table 2's primary basis (Supplementary Note S1.10b, eTable 21); reported here as the disclosed sensitivity comparison, not as a co-primary result (Results, Sensitivity Analyses). Unlike Table 2, this table includes the uniform-prior row, for which a calibrated basis is not defined. Accuracy under the uniform prior is, selection by selection, identical to neural-only accuracy, because a uniform prior leaves the neural posterior unchanged; the two tables in the main text nonetheless display different values (71.6% here vs 71.8% in Table 1) because this table's accuracy is participant-weighted and Table 1's is selection-weighted (Table 1 note). Under the uniform prior, neural contribution fraction, phantom agreement, and prior capture are fixed by construction, because a uniform prior cannot displace the neural posterior, so no CI is shown for those three measures in that row; accuracy still varies across the participant-cluster bootstrap and so retains a genuine CI.

| Prior | Parameters | Neural contribution fraction (95% CI) | Phantom agreement, % (95% CI) | Prior capture, % (95% CI) | Accuracy, % (95% CI) |
| --- | --- | --- | --- | --- | --- |
| Uniform | None | 1.000 | 0.0 | 0.00 | 71.6 (66.1 to 77.2) |
| Character 5-gram | None | 0.866 (0.841 to 0.892) | 8.6 (7.0 to 10.1) | 0.75 (0.42 to 1.03) | 79.5 (75.0 to 84.1) |
| 5-gram, Kneser-Ney | None | 0.885 (0.860 to 0.909) | 7.7 (6.1 to 9.2) | 0.46 (0.27 to 0.63) | 78.9 (74.4 to 83.4) |
| 5-gram, Kneser-Ney (WikiText-103) | None | 0.900 (0.878 to 0.923) | 6.9 (5.4 to 8.4) | 0.39 (0.20 to 0.56) | 78.2 (73.6 to 82.9) |
| GPT-2 | 124M | 0.939 (0.924 to 0.955) | 3.4 (2.6 to 4.1) | 1.55 (1.03 to 2.02) | 73.5 (68.2 to 79.0) |
| GPT-2 large | 774M | 0.937 (0.922 to 0.953) | 3.5 (2.8 to 4.2) | 1.66 (1.14 to 2.13) | 73.5 (68.1 to 79.0) |
| Qwen2.5-1.5B | 1.54B | 0.925 (0.908 to 0.944) | 4.1 (3.3 to 4.8) | 1.65 (1.21 to 2.04) | 74.1 (68.9 to 79.4) |
| Qwen2.5-3B | 3.09B | 0.927 (0.910 to 0.945) | 3.1 (2.5 to 3.7) | 2.55 (1.87 to 3.13) | 72.2 (66.9 to 77.7) |
| Qwen2.5-14B | 14.7B | 0.923 (0.905 to 0.942) | 3.9 (3.1 to 4.6) | 1.83 (1.29 to 2.33) | 73.7 (68.4 to 79.1) |
| Qwen3.5-27B (Instruct) | 27.0B | 0.940 (0.925 to 0.955) | 3.2 (2.5 to 3.8) | 1.31 (0.89 to 1.69) | 73.5 (68.3 to 78.9) |
| Qwen2.5-32B | 32.5B | 0.925 (0.907 to 0.943) | 3.8 (3.0 to 4.5) | 1.80 (1.29 to 2.27) | 73.6 (68.4 to 79.1) |
| Qwen3.6-35B-A3B (Instruct) | 35.0B | 0.932 (0.917 to 0.949) | 3.7 (2.9 to 4.4) | 1.25 (0.84 to 1.60) | 74.1 (68.9 to 79.3) |
| Llama-3.1-8B (Instruct) | 8.0B | 0.927 (0.910 to 0.945) | 3.9 (3.2 to 4.6) | 1.90 (1.24 to 2.46) | 73.6 (68.4 to 79.0) |
| Gemma-2-2B (Instruct) | 2.0B | 0.916 (0.897 to 0.936) | 3.0 (2.5 to 3.6) | 3.31 (2.67 to 3.95) | 71.4 (65.8 to 77.0) |
| Gemma-2-9B | 9.0B | 0.907 (0.888 to 0.927) | 4.9 (3.8 to 5.9) | 2.93 (2.36 to 3.47) | 73.6 (68.5 to 78.8) |
| Gemma-2-27B | 27.0B | 0.944 (0.930 to 0.959) | 3.2 (2.4 to 3.9) | 1.59 (1.15 to 1.99) | 73.3 (68.0 to 78.7) |
| Gemma-4-12B (Instruct) | 12.0B | 0.931 (0.915 to 0.948) | 2.5 (2.0 to 2.9) | 2.46 (1.63 to 3.18) | 71.7 (66.2 to 77.2) |
| Mistral-7B | 7.0B | 0.936 (0.922 to 0.952) | 4.2 (3.4 to 4.9) | 1.64 (1.10 to 2.12) | 74.2 (69.0 to 79.5) |
| Mixtral-8x7B | 46.7B | 0.933 (0.918 to 0.949) | 4.2 (3.4 to 5.0) | 1.77 (1.26 to 2.22) | 74.1 (68.9 to 79.5) |
| DeepSeek-V2-Lite | 15.7B | 0.930 (0.913 to 0.947) | 4.2 (3.4 to 5.0) | 1.87 (1.40 to 2.32) | 74.0 (68.9 to 79.4) |
| OLMo-2-7B (Instruct) | 7.0B | 0.908 (0.887 to 0.929) | 4.1 (3.3 to 4.9) | 2.48 (1.69 to 3.17) | 73.2 (68.2 to 78.5) |
| GPT-OSS-20B (Instruct) | 20.9B | 0.936 (0.921 to 0.952) | 3.2 (2.5 to 3.9) | 1.88 (1.34 to 2.36) | 73.0 (67.7 to 78.4) |
| Pythia-12B | 12.0B | 0.926 (0.910 to 0.943) | 4.0 (3.2 to 4.8) | 1.72 (1.16 to 2.22) | 73.9 (68.7 to 79.4) |
| Mamba-2.8B | 2.8B | 0.929 (0.913 to 0.946) | 3.8 (3.1 to 4.5) | 1.79 (1.29 to 2.21) | 73.7 (68.5 to 79.1) |
| RecurrentGemma-2B | 2.7B | 0.944 (0.930 to 0.959) | 3.4 (2.7 to 4.1) | 1.36 (0.95 to 1.76) | 73.7 (68.4 to 79.1) |

**eTable 23. No monotonic association with parameter count or architecture family was detected, calibrated fusion (primary basis).** The calibrated-basis counterpart of eTable 9, computed on the held-out, per-source-calibrated ladder (Supplementary Note S1.10b, eTable 21) and reported in the Results as the primary basis for the Effect of Parameter Count section. Same construction as eTable 9 otherwise: Spearman rank correlations between each attribution measure and the base-10 logarithm of parameter count, across the 21 neural language models spanning eleven architecture families, with the final two columns repeating each correlation on the ladder truncated below 10 billion parameters (11 of the 21 neural language models).

| Measure | Range across the ladder | rho, full ladder | P, full ladder | rho, truncated <10B | P, truncated <10B |
| --- | --- | --- | --- | --- | --- |
| Phantom agreement | 2.1 to 5.2% | -0.03 | .91 | +0.14 | .68 |
| Neural contribution fraction | 0.895 to 0.971 | -0.04 | .86 | +0.01 | .97 |
| Prior capture | 1.7 to 3.0% | -0.08 | .74 | +0.09 | .79 |
| Accuracy gained by adding the prior | 0.4 to 2.4 points | +0.18 | .45 | +0.31 | .36 |

All four full-ladder correlations remain null under calibration, as they were on the raw basis. eTable 9's one truncated-range exception, phantom agreement's marginal correlation among the 11 priors below 10 billion parameters (rho = +0.62; *P* = .044, raw basis), does not reproduce here (rho = +0.14; *P* = .68): under calibration the truncated-range signal disappears entirely rather than merely attenuating, consistent with the raw finding being a property of the sampled range rather than a real parameter-count effect.

**eTable 24. Symmetric pre- and post-scaling calibration diagnostics for both sources (Supplementary Note S1.13).** Mean negative log-likelihood of the intended symbol, Brier score (full 36-way one-hot target, 0-to-2 scale), and top-label expected calibration error (confidence = max probability, correctness = arg max matches the intended symbol; 10 equal-width bins), computed independently for the neural posterior and the language-model prior, before scaling (raw posteriors, T = 1) and after scaling (the same held-out, 5-fold per-source temperature calibration underlying eTable 21 and this study's principal co-primary estimates). All rows n = 3,373 selections at the primary prior (Qwen2.5-32B).

| Source | Basis | NLL | Brier, 0-2 | ECE, % |
| --- | --- | --- | --- | --- |
| Neural posterior | Pre-scaling (T = 1) | 2.091 | 0.438 | 16.7 |
| Neural posterior | Post-scaling (held-out T) | 1.189 | 0.392 | 4.8 |
| Language-model prior | Pre-scaling (T = 1) | 3.836 | 1.111 | 34.7 |
| Language-model prior | Post-scaling (held-out T) | 3.253 | 0.951 | 4.1 |

Every measure, for both sources, improved after scaling. The language-model prior was the more severely miscalibrated of the two sources pre-scaling (34.7% versus 16.7% ECE) despite requiring a smaller held-out temperature correction than the neural posterior (T approximately 2.4 versus approximately 3.1, eTable 21); post-scaling the two sources converge to a similar residual ECE (4.1 to 4.8%), which is not zero and is not interpreted here as evidence that either source is calibrated in an absolute sense.

**eTable 25. Next-character predictive quality is associated with attribution among neural language models (validity check restricted to the 21 neural language models).** eTable 9 and eTable 23 test whether attribution scales with a prior's parameter count and detect no monotonic association, on either the raw or calibrated basis. This table asks a related but distinct question: does attribution scale with a prior's own measured predictive quality, independent of its size? Predictive quality is each prior's mean negative log-likelihood of the intended character, a task-specific property of the model under the character-projection interface used in this study (Methods), evaluated on the same 3,373 study selections used throughout, rather than a property of how it fuses with the neural decoder. Spearman rank correlations are computed against phantom agreement and neural contribution fraction, both under the held-out, per-source-calibrated fusion of S1.10b/eTable 21 (the study's primary basis), restricted to the 21 neural language models. The 3 character n-gram variants (eTable 19) have a well-defined next-character NLL, computed on the same study selections in the same way, and they are plotted on Figure 3 panel a's NLL axis alongside the 21; they are excluded from this correlation so that it measures how attribution tracks predictive quality within a single model class rather than partly reflecting the difference between classes. The n-grams are character-native models fit by frequency smoothing over a fixed corpus, whereas the 21 causal-LM priors are pretrained general-purpose checkpoints whose next-character distribution is marginalized from a subword-tokenized one (Methods). The correlation across all 24 priors is reported descriptively in the Figure 3 caption.

| Comparison | n | rho | P |
| --- | --- | --- | --- |
| Phantom agreement vs. NLL | 21 | -0.64 | .002 |
| Neural contribution fraction vs. NLL | 21 | +0.75 | <.001 |

Both correlations run in the same direction: priors with lower (better) NLL supply a larger share of the fused posterior's displacement (lower neural contribution fraction) and more often supply the specific evidence that recovers a selection the neural decoder alone would have missed (higher phantom agreement). This is not in tension with the parameter-count null result above: eTable 9 and eTable 23 test whether raw pretraining scale predicts attribution and find that it does not; this table tests whether a prior's own measured task-specific prediction quality predicts attribution, a different axis, on which it does. Read together, the two results indicate the attribution framework tracks a prior's actual next-character predictive competence rather than a proxy such as parameter count or architecture family.

**eTable 26. Neural support among phantom-agreement selections, calibrated fusion (primary basis).** The calibrated-basis counterpart of eTable 11, computed on the held-out, per-source-calibrated fusion of S1.10b/eTable 21 that is Figure 4's own basis and the primary basis for the Results paragraph beside it. For every selection where the calibrated fused posterior selected the intended character but the calibrated neural posterior alone would not have (n = 166), the neural probability assigned to the intended character and its rank within the 36-symbol neural posterior.

| Neural probability on intended character | Share of phantom-agreement cases |
| --- | --- |
| Below 0.10 | 24.1% |
| 0.10 to 0.40 | 71.7% |
| 0.40 or above | 4.2% |

| Neural rank of intended character | Share of all 166 cases |
| --- | --- |
| Second-ranked (neural's runner-up) | 59.0% |
| Third-ranked | 21.1% |
| Fourth-ranked or worse | 17.5% |
| Tied with neural's top symbol | 2.4% |

Median neural probability on the intended character: 0.163. Median margin between the neural posterior's own top choice and the intended character: 0.108. The tied row (4 cases, 2.4%) is where the intended character matched the neural posterior's own top-probability symbol exactly, shown separately from the three ranked rows rather than folded into "second-ranked"; a phantom-agreement label is still possible in this situation because the deterministic tie-break used to convert the neural posterior into a single decoded symbol did not resolve to the intended character even though its probability was not exceeded by any other symbol.

**eTable 27. Closed-loop emitted-context sensitivity for the primary prior, calibrated fusion, paired comparison (primary basis) (Supplementary Note S1.14).** The calibrated-basis counterpart of eTable 16, computed on the held-out, per-source-calibrated fusion of S1.10b/eTable 21 that is the primary basis for the Results paragraph beside it. Unlike eTable 16's two independently weighted and resampled rows, this table reports one paired participant-cluster bootstrap difference (emitted minus intended context) per measure: both arms share the same per-fold calibration temperatures, already fit on the intended-context data (S1.10b), so the two sides differ only in which context they score, not in how they are calibrated. The second row instead independently refits a temperature on the emitted arm, a labeled sensitivity comparison rather than the primary basis.

| Comparison | Neural contribution fraction difference (emitted minus intended) | 95% CI | Phantom agreement difference (emitted minus intended), pp | 95% CI |
| --- | --- | --- | --- | --- |
| Shared temperature (primary) | -0.019 | -0.025 to -0.013 | -0.7 | -1.4 to -0.1 |
| Independently recalibrated (sensitivity) | -0.007 | -0.012 to -0.001 | -0.8 | -1.5 to -0.2 |

n = 3,373 for both rows. Both rows found emitted (closed-loop) context selections had a lower neural contribution fraction and lower phantom agreement than the same selections' intended context, each 95% CI excluding 0, the same divergent pattern already reported on the raw basis (eTable 16): a larger prior share paired with fewer phantom-agreement corrections, not the uniformly weaker attribution the permutation negative control shows (eTable 15, eTable 29). Independently refitting the temperature on the emitted arm attenuated the neural-contribution- fraction gap roughly threefold (-0.019 to -0.007) without materially changing the phantom-agreement gap, consistent with the calibrator partially absorbing part of the context-drift effect under study when fit on the very data whose drift is being measured; the shared-temperature row is reported as the primary comparison for that reason. Both rows are paired comparisons: the same 3,373 selections' calibrated intended- and emitted-context estimates are differenced within each participant-cluster bootstrap replicate, so, unlike eTable 16, the two arms are on identical statistical footing by construction.

**eTable 28. Position, decoder quality, and target type, calibrated fusion (primary basis for Secondary Outcomes) (sensitivity comparison to eTable 2, eTable 3, eTable 4, and eTable 20).** The calibrated-basis counterpart of eTable 2 (position), the attribution columns of eTable 3 (decoder quality; reconstruction fidelity itself has no calibrated analog), and eTable 4 (target type), computed on the held-out, per-source-calibrated fusion of S1.10b/eTable 21 that is the primary basis for the Results' Secondary Outcomes section. Estimates are participant-weighted with 95% CIs from 2,000 participant-cluster bootstrap replicates.

| Position | Selections | Phantom agreement, % | 95% CI | Neural contribution fraction | 95% CI |
| --- | --- | --- | --- | --- | --- |
| 1 | 662 | 2.5 | 1.6, 3.3 | 0.934 | 0.918, 0.951 |
| 2 | 662 | 4.2 | 2.6, 5.6 | 0.926 | 0.909, 0.944 |
| 3 | 661 | 3.8 | 2.6, 4.9 | 0.909 | 0.890, 0.930 |
| 4 | 542 | 4.2 | 2.8, 5.4 | 0.903 | 0.882, 0.925 |
| 5 | 429 | 9.2 | 6.4, 11.6 | 0.869 | 0.840, 0.898 |
| 6 | 417 | 7.7 | 5.3, 10.0 | 0.879 | 0.854, 0.903 |

The position gradient's robustness to word-identity and session confounding (eTable 20) also holds under calibration. These three coefficients are the linear mixed-model sensitivity comparison's own, the model eTable 20's neural refits are built on: the unadjusted calibrated coefficient is -0.0081 (95% CI, -0.0103 to -0.0058; *P* < .001), the word- and session-adjusted coefficient -0.0082 (95% CI, -0.0103 to -0.0062; *P* < .001), and the six-character-words-only coefficient -0.0085 (95% CI, -0.0110 to -0.0059; *P* < .001), each close to the others, as on the raw basis.

Phantom agreement's own position gradient, the 2.5% to 7.7% rise across the six positions in the table above, holds under the same adjustment on this basis: the unadjusted calibrated odds ratio per character is 1.19 (95% CI, 1.07 to 1.32; *P* = .001, Benjamini-Hochberg-adjusted *P* = .002; eTable 30), and the word- and session-adjusted calibrated odds ratio is 1.17 (95% credible interval, 1.11 to 1.23). As on the raw basis, the adjusted estimate comes from a crossed-random-effects Bayesian mixed generalized linear model fit by variational approximation, so its interval is a credible interval rather than a confidence interval, no P value is defined for it, and it is not a member of the Benjamini-Hochberg family (Supplementary Note S1.6b, eTable 20). That model's random-effect standard deviations, on the log-odds scale, are 0.45 for participant (95% credible interval, 0.37 to 0.56), 0.46 for word identity (0.42 to 0.50), and 0.20 for session (0.18 to 0.23), participant and word identity being comparable in size here rather than word identity dominating as on the raw basis. No six-character-words-only refit is reported for phantom agreement on either basis.

| Tertile | Selections | Phantom agreement, % | 95% CI | Neural contribution fraction | 95% CI |
| --- | --- | --- | --- | --- | --- |
| Low | 1,165 | 7.2 | 5.6, 8.6 | 0.853 | 0.821, 0.887 |
| Middle | 1,087 | 3.9 | 2.5, 5.3 | 0.941 | 0.921, 0.962 |
| High | 1,121 | 2.7 | 1.5, 3.7 | 0.959 | 0.946, 0.973 |

| Target type | Selections | Phantom agreement, % | 95% CI | Neural contribution fraction | 95% CI |
| --- | --- | --- | --- | --- | --- |
| English word | 3,205 | 4.3 | 3.5, 5.2 | 0.915 | 0.897, 0.934 |
| Digit string | 168 | 6.6 | 2.2, 10.3 | 0.885 | 0.830, 0.947 |

Every stratum's calibrated phantom agreement is higher, and its calibrated neural contribution fraction lower, than its raw-basis counterpart in eTable 2, eTable 3, and eTable 4, the same directional shift the co-primary estimates show relative to their own raw-fusion sensitivity comparison (Results). The corresponding hypothesis tests, with Benjamini-Hochberg-adjusted P values, are reported in eTable 30.

**eTable 29. Stratified context-permutation negative control, calibrated fusion (primary basis) (sensitivity comparison to eTable 15) (Supplementary Note S1.12).** The calibrated-basis counterpart of eTable 15, computed on the held-out, per-source-calibrated fusion of S1.10b/eTable 21; per-fold calibration temperatures were fit once on the real (unpermuted) permutable subset only and applied identically to each selection's real evaluation and to every permuted draw involving it, never refit per permutation draw or on permuted data. Values are unweighted means, as in eTable 15.

| Context | Neural contribution fraction | Phantom agreement |
| --- | --- | --- |
| Real (true, matched) context | 0.897 | 5.6% |
| Permutation null, mean across 200 permutations (95% CI) | 0.901 (0.899 to 0.902) | 3.3% (3.0 to 3.7%) |

The real context's phantom agreement (5.6%) exceeded every one of the 200 permutation draws (one-sided exceedance *P* < .005) and its neural contribution fraction (0.897) fell below every draw (*P* < .005), the same direction and conclusion as the raw-basis check (eTable 15): a real but wrong context supplies less useful evidence than the true context, weighing against an alternative explanation in which any sufficiently confident prior, regardless of relevance, would produce the same phantom-agreement rate.

**eTable 30. Secondary-family hypothesis tests with multiplicity correction, calibrated fusion (primary basis) (sensitivity comparison to eTable 5).** The calibrated-basis counterpart of eTable 5, same five-member family and Benjamini-Hochberg procedure, evaluated on the calibrated models built alongside the held-out, per-source-calibrated fusion of S1.10b/eTable 21. The first two neural contribution fraction rows are the primary effect estimates for the two continuous relationships: predicted-value contrasts from the bounded fractional-logit model, with delta-method 95% CIs on the participant-clustered sandwich covariance (Supplementary Note S1.6). They are reported outside the five-member Benjamini-Hochberg family, whose continuous members are the linear mixed-model slopes in the last two rows; that model is retained as the labeled sensitivity comparison and is the source of this outcome's adjusted P values.

| Test | Estimate | 95% CI | Raw P | Adjusted P |
| --- | --- | --- | --- | --- |
| Phantom agreement per character of context, odds ratio | 1.19 | 1.07, 1.32 | 0.001 | 0.002 |
| Phantom agreement per unit calibration AUC, odds ratio | 0.030 | 0.005, 0.204 | <0.001 | <0.001 |
| Phantom agreement, digit versus word target, odds ratio | 1.45 | 0.70, 3.02 | 0.323 | 0.323 |
| Neural contribution fraction across word positions 1 to 6, bounded-model predicted contrast | -0.039 | -0.050, -0.027 | <0.001 | Not in family |
| Neural contribution fraction between the outer tertiles' median calibration AUC, bounded-model predicted contrast | +0.082 | +0.056, +0.109 | <0.001 | Not in family |
| Linear mixed-model sensitivity comparison: neural contribution fraction per character of context, slope | -0.0081 | -0.0103, -0.0058 | <0.001 | <0.001 |
| Linear mixed-model sensitivity comparison: neural contribution fraction per unit calibration AUC, slope | +0.442 | +0.373, +0.511 | <0.001 | <0.001 |

Every test's conclusion matches its raw-basis counterpart (eTable 5): the digit-versus-word comparison is the only one of the five family members not meeting the adjusted threshold, on either basis. Both bounded contrasts agree in direction and significance with the linear slopes they replace as the primary summary.

**eTable 31. Formal tests of the parameter-count and architecture-family null result, calibrated fusion (primary basis) (sensitivity comparison to eTable 18) (Supplementary Note S1.16).** The calibrated-basis counterpart of eTable 18: a permutation test for whether architecture family explains variance in each attribution measure beyond chance, and a two-one-sided-tests (TOST) equivalence test for the Spearman correlation between log parameter count and each measure against a predefined margin of rho = ±0.3, both computed on the calibrated ladder (Table 2, eTable 23).

| Measure | Permutation F-like statistic | Permutation *P* | Spearman rho | 90% CI | Equivalent at ±0.3? |
| --- | --- | --- | --- | --- | --- |
| Neural contribution fraction | 1.95 | .18 | -0.04 | -0.44 to 0.33 | No |
| Phantom agreement | 1.07 | .45 | -0.03 | -0.39 to 0.37 | No |
| Prior capture | 2.01 | .12 | -0.08 | -0.46 to 0.34 | No |
| Accuracy gained by adding the prior | 0.92 | .53 | +0.18 | -0.19 to 0.49 | No |

Neither test found an architecture-family or parameter-count effect on the calibrated basis that the raw-basis tests had not already suggested (eTable 18): no measure's between-family variance exceeded chance, and no measure's equivalence-test CI fell entirely within the predefined margin. The calibrated-basis CIs are wider than their raw-basis counterparts for every measure, consistent with the calibrated ladder carrying more between-fold variability than the raw one; as on the raw basis, these results are better read as a failure to reject a parameter-count or architecture-family effect than as positive evidence that none exists.

**eTable 32. Leave-one-study-out sensitivity, calibrated fusion (primary basis) (sensitivity comparison to eTable 7).** The calibrated-basis counterpart of eTable 7: each row removes one source study and recomputes both co-primary outcomes, under the held-out, per-source-calibrated fusion of S1.10b/eTable 21, on the remainder.

| Study removed | Phantom agreement, % | Neural contribution fraction |
| --- | --- | --- |
| Study B | 6.3 | 0.880 |
| Study F | 4.2 | 0.913 |
| Study L | 4.1 | 0.921 |
| Study N | 3.5 | 0.935 |

Removing any single source study left calibrated phantom agreement between 3.5% and 6.3% and the neural contribution fraction between 0.880 and 0.935, the range the main text reports (Results, Sensitivity Analyses); the ordering across studies matches eTable 7's raw-basis counterpart.

### S3 Supplementary Figures

**eFigure 1. Both co-primary outcomes vary with the fusion exponent, monotonically on the raw basis and closely, but not exactly, so on the calibrated basis.** (a) Phantom agreement and (b) neural contribution fraction, raw, uncalibrated fusion at each of the five prespecified exponents; (c) and (d) the same two outcomes re-fused from the held-out, per-source calibrated posteriors of Figure 1's co-primary estimates and of Table 2. The same 3,373 selections from 47 participants with ALS are re-fused at every exponent, on each basis in turn. The x-axis is log2-scaled: the five prespecified exponents (0.25, 0.5, 1, 2, 4) are exactly 2^-2^ through 2^2^, so base-2 spacing places every value at an integer power and all five land evenly spaced with genuine numerical meaning; a log10 axis, used for an earlier version of this panel, clustered four of the five values between decade ticks and labeled only one. Points are participant-weighted means: the filled point at beta = 1 is the prespecified exponent, the value every estimate reported in this study uses, and the four open points are sensitivity-only exponents, at which no other result is computed. Bars are 95% CIs from 2,000 participant-cluster bootstrap replicates over the 47 participants; the interval at beta = 1 in (a) and (b) is the raw-fusion co-primary interval the main text reports as a sensitivity comparison (phantom agreement 3.8%, 95% CI 3.0 to 4.5; neural contribution fraction 0.925, 95% CI 0.907 to 0.943), and the interval at beta = 1 in (c) and (d) is the calibrated co-primary interval the main text reports as the principal result (phantom agreement 4.4%, 95% CI 3.5 to 5.3; neural contribution fraction 0.914, 95% CI 0.896 to 0.934). (a) and (b) are computed on raw, uncalibrated posteriors at the primary prior (Qwen2.5-32B); raw beta = 1 fusion remains the basis only for a specific raw-only minority of analyses (the alternative attribution formalisms), alongside this exponent grid and the temperature-scaling grid, which vary those parameters by design; every other secondary and sensitivity analysis is calibrated-primary. On the raw basis, the direction of every finding is monotonic across the full grid. On the calibrated basis, phantom agreement rises slightly from beta = 0.25 to beta = 0.5 (5.4% to 5.8%) before falling, and neural contribution fraction peaks at beta = 2 (0.916) rather than at beta = 4 (0.898); both deviations sit within overlapping 95% CIs. At the lowest calibrated exponent (beta = 0.25) specifically, neural contribution fraction falls to 0.101 and prior capture rises to 21.6% (eTable 21), a reversal, at that sensitivity-only exponent alone, of the finding that neural evidence supplies most of the KL-attributed posterior displacement at the prespecified beta = 1 the co-primary estimates use; calibration raises the fitted neural temperature well above 1 (eTable 21), so down-weighting an already-flattened neural posterior further at beta < 1 hands most selections to the prior. The magnitude of phantom agreement varies from 1.4% to 5.7% raw and from 1.7% to 5.8% calibrated.

**eFigure 2. Phantom agreement by source study and stimulation condition.** One panel per source study and one row per condition that study ran. Points are participant-weighted means and bars are 95% CIs from 2,000 participant-cluster bootstrap replicates over the 8 to 18 participants contributing to each row. Point area increases with the number of participants behind the estimate, the bootstrap's clustering unit, and the legend below the panels gives the area at three reference counts (8, 13, 18 participants); because conditions within a study share the same participants, all rows in a panel share one point size. The number of selections behind each estimate is printed separately at the right of each row and ranges from 234 to 846; the dashed vertical line is the whole-cohort estimate of 4.4%, the calibrated co-primary the main text reports as the principal result. Condition names are those of eTable 8, which reports the raw-basis estimates pooled across studies. Conditions are nested within studies in this archive, so most study-by-condition combinations do not exist; only the 9 that do are drawn, in place of the earlier matrix in which 23 of 32 cells were empty and the sample behind each shaded cell was not shown. Together, those 9 combinations span all 3,373 selections from 47 participants with ALS. Estimates are computed on the calibrated-fusion basis (Methods) at the primary prior (Qwen2.5-32B, beta = 1), the basis of Figure 1's co-primary estimates and of Table 2, not the raw, uncalibrated fusion eTable 8 reports as a disclosed sensitivity comparison.

**eFigure 3. The per-selection distribution of the prior's share of posterior displacement is highly right-skewed on both bases, and calibration shifts it rightward.** Empirical cumulative distribution functions of 1 minus the neural contribution fraction across the 3,373 selections from 47 participants with ALS at the primary prior (Qwen2.5-32B, beta = 1): raw, uncalibrated fusion (vermilion), the same quantity and basis as eTable 14's Kullback-Leibler-based measure, and the held-out, per-source calibrated fusion (blue) of Figure 1's co-primary estimates and of Table 2. Both curves are unweighted across selections. The raw mean (0.080) and median (0.001) are marked, and so are the calibrated mean (0.090) and median (0.029); the participant-weighted mean of each -- 7.5% raw, the value the main text reports as a sensitivity comparison, and 8.6% calibrated, the co-primary complement the main text reports as the principal result -- differs from its own curve's unweighted mean by less than half a percentage point and is stated beside that mean mark rather than drawn as a further, indistinguishable line. Because both distributions pile up against zero, the inset redraws the region from 0 to 0.10 at ten times the scale, where 80% of raw selections and 70% of calibrated selections fall, with each median annotated directly; the two-row strip beneath the axis places one tick at each percentile of each distribution, so that the density of ticks is the density of selections, on each basis in its own row. Open markers on each curve, and the table at the upper right, give the fraction of selections below 0.01, 0.05, 0.10 and 0.50 (63%, 74%, 80% and 95% raw; 40%, 57%, 70% and 98% calibrated). Calibration moves mass out of the near-zero pile the raw curve shows and toward larger prior shares across most of the distribution, consistent with the fitted neural temperature well above 1 that eTable 21 reports: a flattened neural posterior hands more of a selection's KL-attributed posterior displacement to the prior, on more selections, than the raw posterior did.

**eFigure 4. No association detected between phantom agreement and parameter count on the raw, uncalibrated ladder.** One point per prior, raw, uncalibrated fusion at the primary prior (Qwen2.5-32B, beta = 1), the same prior as Figure 1's co-primary estimates and the same basis as eTable 22: circles are neural language models, filled when instruction-tuned (eTable 19) and hollow when base; diamonds are character 5-grams and the square is the uniform null, neither of which has a parameter count and so both sit in the left gutter rather than on the log-parameter axis; colour is architecture family. Figure 3 panel a relates phantom agreement to each prior's own next-character prediction quality instead (eTable 25); parameter count did not predict phantom agreement on either basis (eTable 9 raw, eTable 23 calibrated). The calibrated-basis analogue of this question, for neural contribution fraction rather than phantom agreement, is reported in Results (Effect of Parameter Count) and eTable 23. Every prior was scored on the same 3,373 selections from 47 participants with ALS; phantom agreement is a participant-weighted mean and parameter counts are total parameters, including the inactive experts of the four sparsely activated models (eTable 19). The Spearman correlation (rho = +0.11; 90% CI, -0.26 to +0.45; P = .63; 21 neural language models, 11 families) is restricted to those 21 neural language models because the 5-grams and the uniform null have no parameter count to correlate against. Figure 3 panel a restricts its own correlation to the same 21 for a different reason: to keep predictive quality from being conflated with model class. Priors placed close together on the parameter-count axis are drawn up to 0.08 of their true parameter count apart, less than a marker width, so that no marker fully hides another.

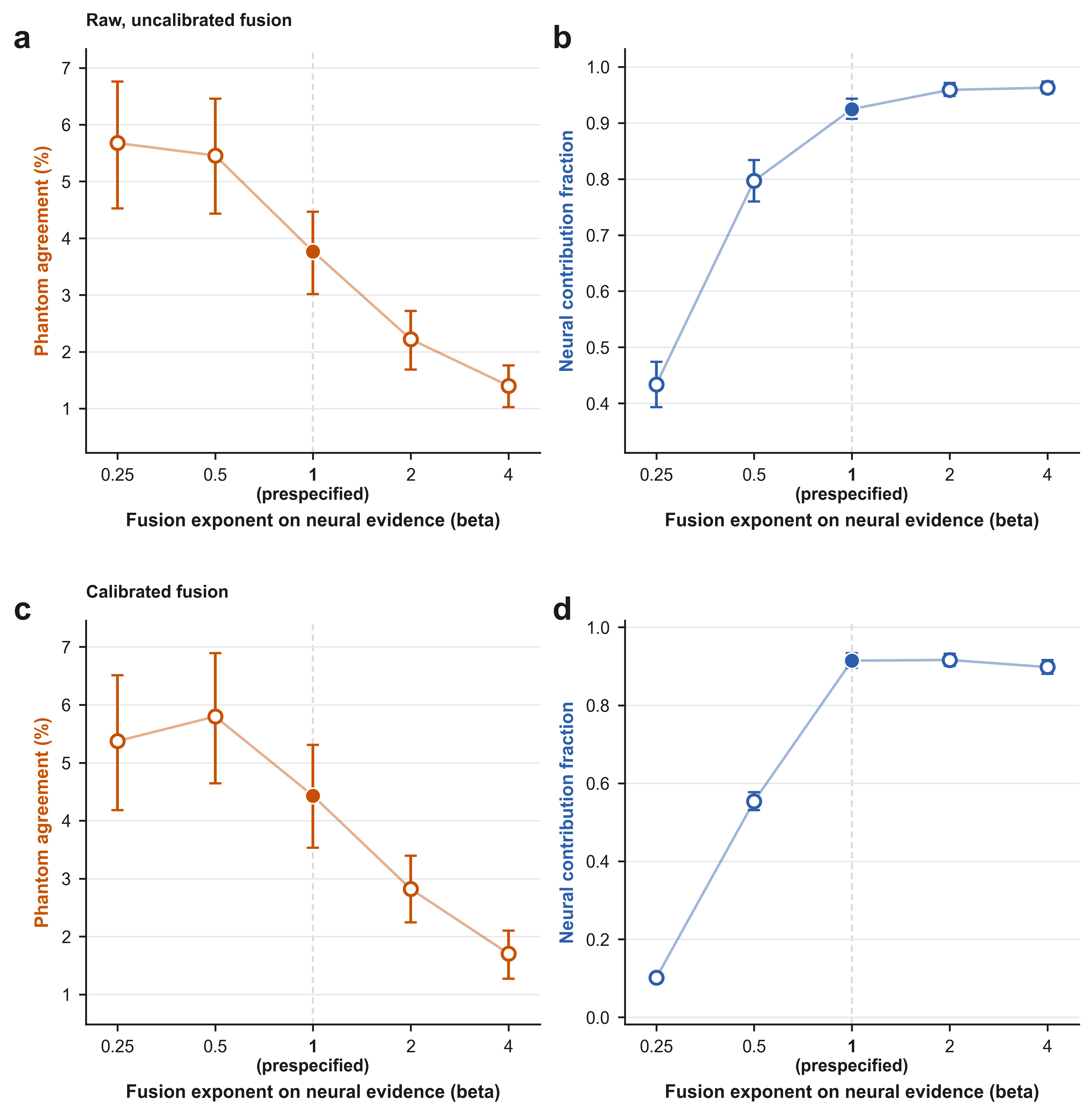

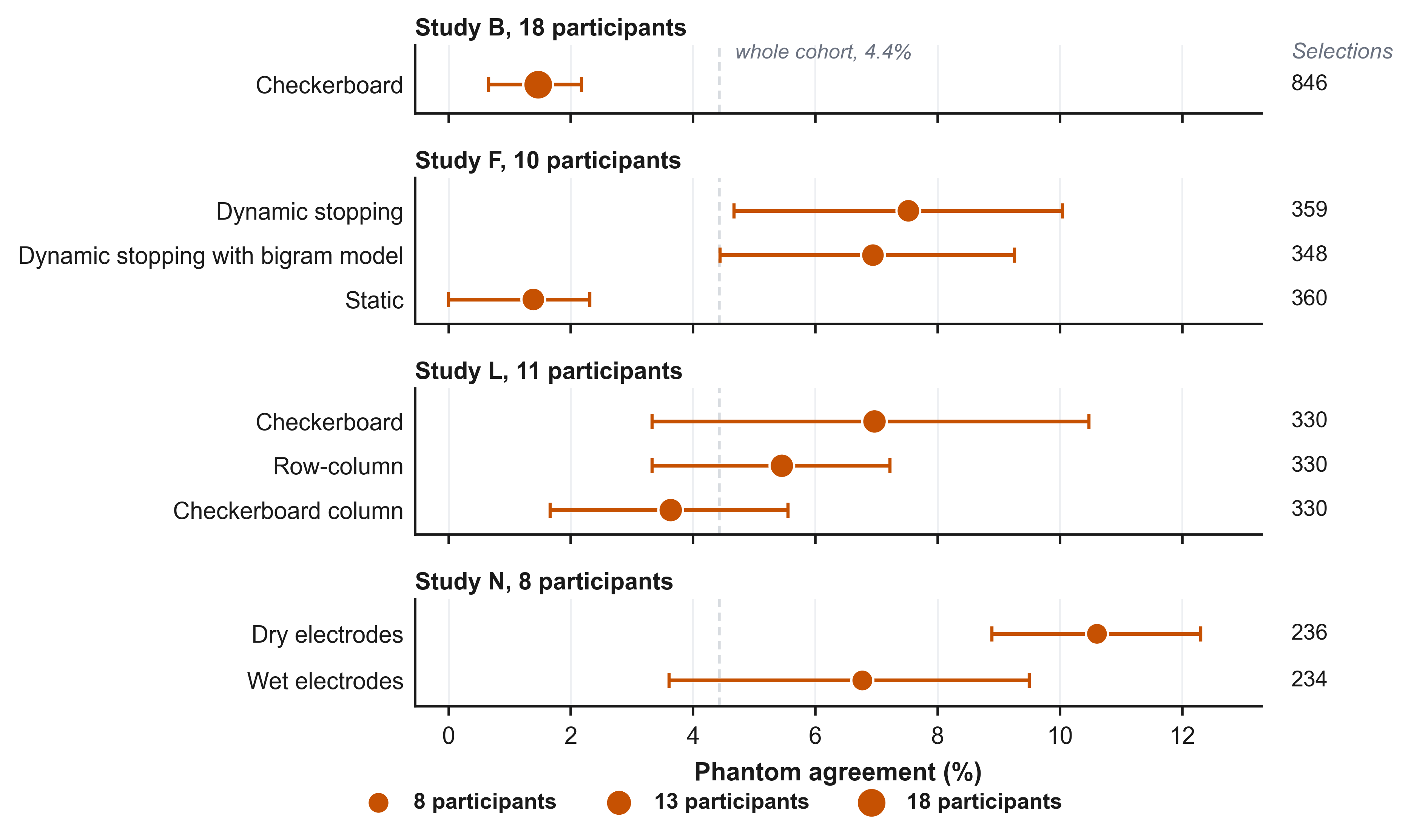

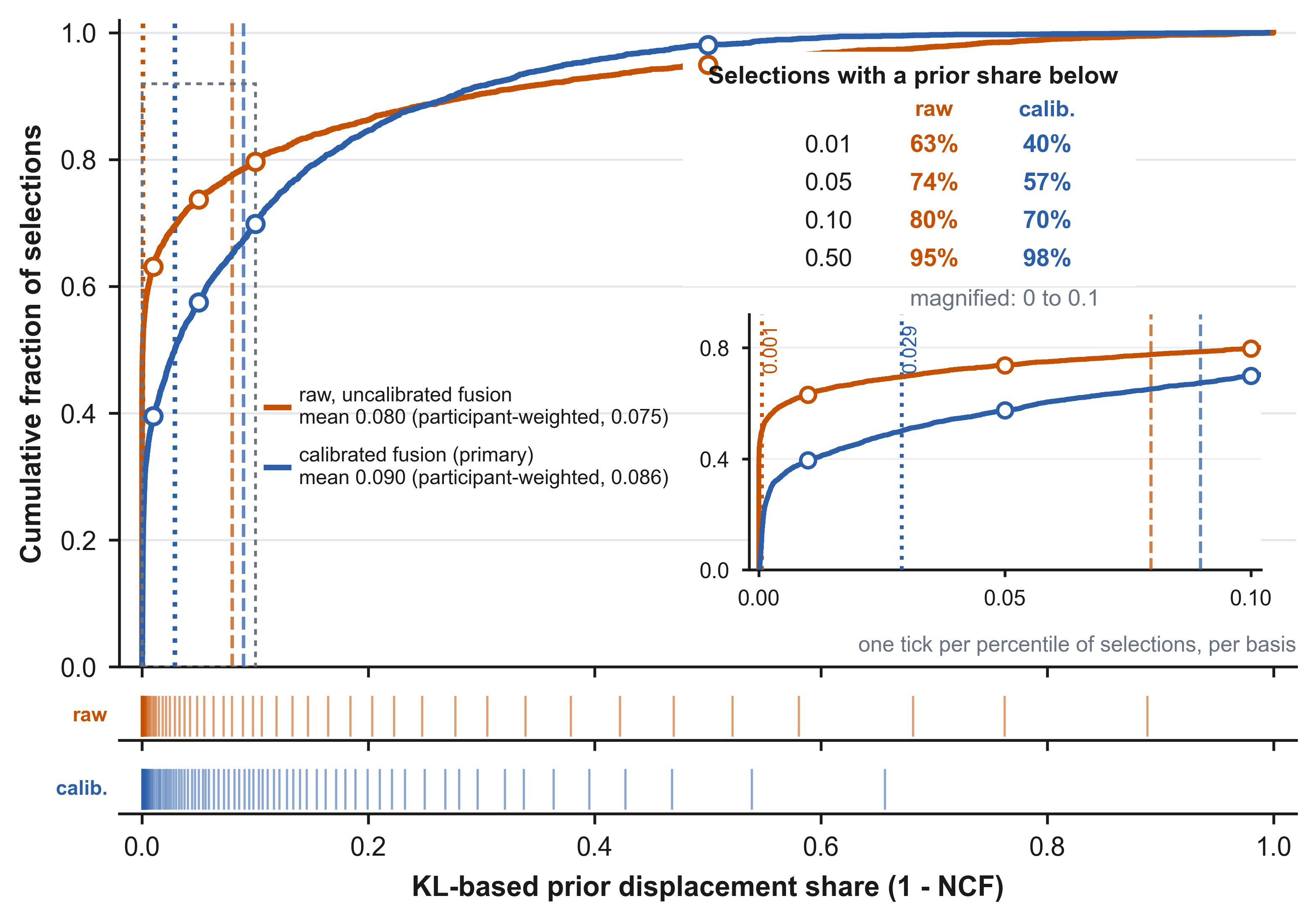

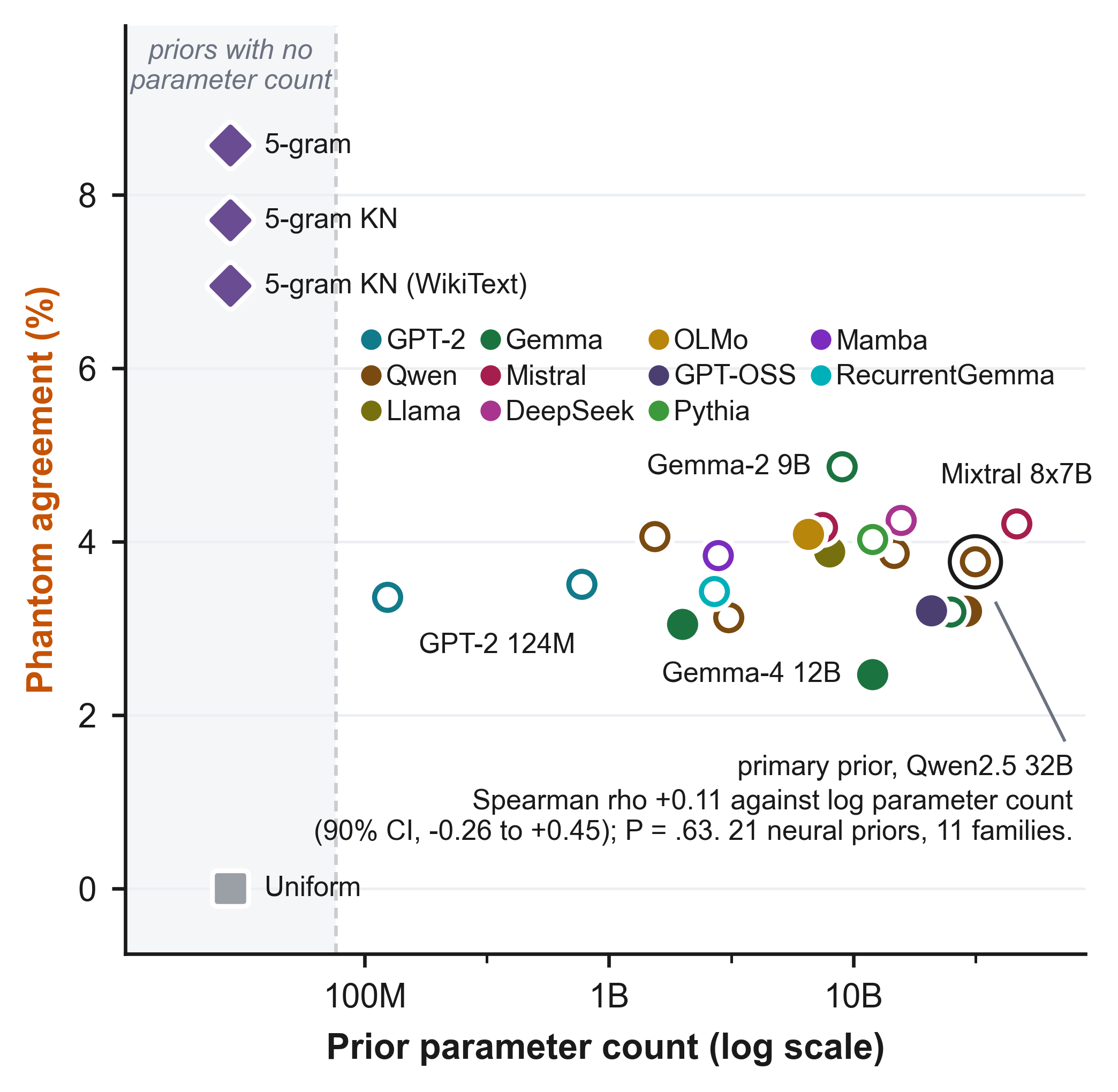

### S4 Code and Data Availability

BigP3BCI version 1.0.0 is publicly available from PhysioNet (doi:10.13026/0byy-ry86). Analysis code, including the attribution formalism, the decoder, the prior ladder, and the statistical pipeline with its regression tests, is publicly available at <https://github.com/BRIDGE-GenAI-Lab/BCI_Authorships>; a permanent archive will be created on Zenodo/OSF upon publication. No individual-level data are redistributed.
